# A Comparative Survey of Functional Evidence Use in Hearing and Vision Loss Genetics

**DOI:** 10.1101/2025.11.25.25339647

**Authors:** R. Arda Inan, Marina T. DiStefano, Sami S. Amr, Tim Beißbarth, Lea M. Starita, Andrew B. Stergachis, Ahmad Abou Tayoun, Robert B Hufnagel, Barbara Vona

## Abstract

**Background:** Advances in sequencing and standardized guidelines for variant classification have improved genetic diagnostics, but a large proportion of variants, particularly in genetically heterogeneous disorders such as hearing and vision loss, remain of uncertain significance. Although emerging multiplexed assays of variant effects (MAVEs) offer a solution to resolve these uncertainties, their integration into clinical practice in these fields remains constrained by limited familiarity and variable implementation among genetics professionals.

**Methods:** We conducted a survey of hearing and ocular genetics professionals (*n* = 82) to explore their practices, experiences, and challenges in variant classification. The survey captured both quantitative and qualitative insights, allowing for an assessment of how experts engage with functional evidence and identifying patterns, perspectives, and areas for potential improvement in clinical workflows.

**Results:** Experts across hearing and vision loss genetics frequently encountered variants of uncertain significance (VUSs) and reported that functional data, while useful, often failed to resolve classification uncertainty due to both limited availability and variable quality of existing studies. Highest confidence was expressed in transcript assays and patient-derived cell models, as well as in computational predictors, while limited confidence or familiarity was reported for non-mammalian models and high-throughput experimental assays. Across both domains, respondents emphasized the need for improved data accessibility, standardized guidelines in the assessment of functional data for different genes or disease areas and expanded training to enhance the integration and clinical utility of functional evidence in variant classification.

**Conclusions:** Our survey identifies both common and domain-specific challenges in applying functional evidence for variant classification in hearing and ocular genetics, emphasizing the need for frameworks that account for biological complexity, methodological limitations, and barriers to data accessibility.

## Background

Sequencing remains crucial for first-line diagnostics in clinical genetics; however, evaluating the clinical impact of the vast number of identified sequence variants represents a significant challenge. Variant classification guidelines have been harmonized by the American College of Medical Genetics and Genomics (ACMG)/Association of Molecular Pathology (AMP) [1], and refined by the Clinical Genome Resource (ClinGen) [2–8], which established Variant and Gene Curation Expert Panels (VCEPs/GCEPs) to provide domain-specific specifications for clinical geneticists. However, despite advances in classification guidelines and the growing availability of genomic and clinical data, the clinical impact of many rare variants remains uncertain. As of July 2025, over 2 million variants, corresponding to 57% of ClinVar entries, are variants of uncertain significance (VUSs) or have conflicting interpretations of pathogenicity [9]. The high abundance of VUS reflects insufficient evidence available to determine clinical significance of these variants, limiting the effectiveness of genetic testing to inform clinical decision-making.

This challenge is particularly evident in genetically heterogeneous disorders such as hearing loss (HL) and vision loss (VL), which share substantial gene overlap. The Genomics England PanelApp lists 191 HL- and 518 VL-associated genes, 41 of which are shared across both panels, as curated by experts [10]. Despite extensive submissions to ClinVar, the burden of VUS remains high, with 44% of HL variants and 46% of VL variants, currently classified as VUS or having conflicting classifications across different clinical labs. The Deafness Variation Database further illustrates the scale of this issue, classifying 89% of variants across 224 deafness-related genes designated as VUS [11].

This VUS prevalence is also reflected in the diagnostic rates of clinical reports, with positive diagnosis in large-scale studies of inherited hearing [12,13] and ocular diseases [14,15], currently between 39-51%. These figures collectively emphasize a challenge that is expected to grow as genomic testing becomes more widespread. and the need for improved variant classification strategies to reduce uncertainty and improve diagnostic accuracy across neurosensory genetic conditions, which affect a substantial proportion of the population.

Multiplexed assays of variant effects (MAVEs) represent a promising, high-throughput avenue for resolving VUS by providing large-scale quantitative functional data in relevant cellular contexts. The field is actively shifting toward proactively creating comprehensive “atlases of variant effects” to reduce uncertainty and enable more accurate variant interpretation [16]. Notably, recent functional studies for *CNGA3* [17] and *KCNQ4* [18] have demonstrated translational utility of medium- and high-throughput assays by contributing to the reclassification of many VUS associated with VL and HL, respectively. While ongoing efforts aim to enable the reclassification of most missense VUSs by 2030 [19], the integration of MAVE data into routine clinical interpretation remains limited, with reports suggesting unclear levels of familiarity, preparedness, and willingness to use it among clinical genetics specialists [20].

A recent survey by Park et al. [21] revealed general challenges faced by genetics professionals regarding the availability, quality, and interpretation of functional data across various fields. Building on these findings, our study focuses specifically on hearing and ocular genetics experts to assess their readiness, expectations, and barriers to adopting functional data in clinical variant classification. We aim to illuminate domain-specific needs and opportunities for integrating functional evidence into specialized diagnostic workflows by concentrating on the common ground shared by these neurosensory fields.

To this end, we conducted a structured survey and received responses from 82 experts from hearing and/or ocular genetics, engaging members of the ClinGen HL VCEP and Ocular Clinical Domain Working Groups. Our goal was to elucidate current challenges in accessing, interpreting, and applying variant-level functional data, particularly in relation to VUS resolution. By fostering cross-domain collaboration, this study seeks to inform future efforts to create resources, guidelines, and collaborations that support the effective use of functional data. Lessons from this effort may also inform broader initiatives to standardize functional evidence interpretation and improve diagnostic yield across rare disease domains.

## Methods

### Survey design and distribution

This survey was inspired by Park et al. [21] but uniquely focused on assessing practices and challenges within the hearing and ocular genetics domains. The questions were carefully developed through a consensus-driven process involving domain experts (A.A.T., S.S.A., M.T.D., R.B.H., and B.V.) via a series of exchanges by email and video conferences. The survey was hosted on a publicly accessible REDCap platform [22] and was open for a period of six months. It was distributed internationally using professional networks of the study team, collecting contacts on PubMed from researching publications reporting on the sequencing of hearing and ocular genetics patient cohorts, through promotion via multiple channels including the ClinGen Resource Quarterly Update, ACMG calls for collaboration, circulation via HL VCEP and Ocular GCEP email lists, Genomics England PanelApp reviewers for hearing loss, auditory neuropathy spectrum disorder, ophthalmological ciliopathies and retinal disorders, social media outreach, and websites or newsletters of affiliated institutions and genetics societies, such as the European Society of Human Genetics. Participation was voluntary, anonymous, and without any direct incentives.

### Survey structure

The survey was conducted in English and consisted of 62 items, including multiple-choice, Likert scale, and open-ended questions on: (1) demographic information, (2) current and future professional activities, (3) scope and challenges of variant interpretation, (4) functional evidence-related tasks, (5) confidence using functional evidence, (6) use of resources that include functional evidence, (7) use of guidelines for application of functional evidence, (8) challenges in utilizing functional evidence, (9) approaches to improving the interaction and application of functional evidence, (10) assessment of existing data on improving clinical utility, (11) use and expectations of gene-level functional data, and (12) use and expectations of variant-level functional data. A complete copy of the survey is provided in **Additional file 1**.

### Survey analysis and visualization

A total of 198 individuals began the survey, resulting in 82 complete responses. While no formal exclusion criteria were applied, individuals who were not actively participating in clinical variant interpretation for either HL or VL were prompted to discontinue the survey. One respondent filled in the physical version of the survey. Their answers were transferred to the REDCap database by authors. Another respondent who only responded to questions related to demographics and professional activities was excluded from further analyses. REDCap data were securely stored on servers of the Society for Scientific Data Processing Göttingen.

All quantitative analyses and visualizations were performed using R (version 4.4.0) with packages including data.table, dplyr, ggplot2, likert, patchwork, reshape2, rnaturalearth, showtext, tidyr, writexl. Median Likert scores were calculated to identify differences across expert groups. Open-ended answers related to handling conflicting functional evidence (n = 37) were analyzed for common thematic elements after filtering out irrelevant answers. Responses to other open-ended questions were discussed contextually within the text where relevant.

### Ethics declaration

The study protocol was reviewed and approved by the Ethics Committee of the University Medical Center Göttingen under protocol number 12/8/24 An.

## Results

### Participant demographics, professional focus and experiences with VUS

We received 82 complete responses to the survey, following direct email contact with 477 participants and broad promotion across professional channels, representing 30 countries, with the highest participation from the United States of America (16%) and Germany (13%) (**Additional file 2: Fig. S1A)**. The majority (73%) worked in academic medical centers and 41% of respondents were affiliated with ClinGen, with varying degrees of experience in variant classification (**Additional file 2: Fig. S1B-C)**. Participants were divided into three main expert groups: 48% (39/82) specialized in hearing loss (HL), 23% (19/82) in vision loss (VL), and 29% (24/82) had cross-disciplinary expertise (H+V). Clinical roles were slightly more prevalent than research roles among HL experts (49% vs 31%), whereas research roles predominated among VL experts (52% vs 37%) (**Table 1**). The respondents represent a diverse and multidisciplinary group, ensuring broad perspectives on variant interpretation in the fields of hearing and ocular disorders.

**Table 1.** Participant characteristics by expert group.

|  | H (n = 39) | H+V (n = 24) | V (n = 19) | Total (n = 82) |
| --- | --- | --- | --- | --- |
| **Sex** |  |  |  |  |
| Male | 36% | 50% | 26% | 38% |
| Female | 56% | 42% | 63% | 54% |
| Not specified | 8% | 8% | 11% | 8% |
| **Leadership role** |  |  |  |  |
| Yes | 49% | 62% | 58% | 55% |
| No | 49% | 38% | 42% | 44% |
| Not specified | 2% | - | - | 1% |
| **Professional position** |  |  |  |  |
| Clinical geneticist | 26% | 17% | 5% | 18% |
| Genetic counselor | 3% | 4% | 16% | 6% |
| Laboratory director | 15% | 25% | 11% | 17% |
| Laboratory technician | 3% | 4% | - | 3% |
| Laboratory medical geneticist | 13% | 21% | 11% | 15% |
| Molecular genetic pathologist | 5% | - | - | 2% |
| Ophthalmologist | - | - | 5% | 1% |
| Pathologist | 2% | - | - | 1% |
| Research scientist | 26% | 12% | 42% | 26% |
| Variant review scientist | 2% | 17% | 10% | 9% |
| Other | 5% | - | - | 2% |
| **Years of professional experience** |  |  |  |  |
| 0-5 | 31% | 21% | 26% | 27% |
| 6-10 | 13% | 25% | 21% | 18% |
| 11-15 | 18% | 25% | 5% | 17% |
| 16-20 | 7% | 8% | 11% | 9% |
| More than 20 | 31% | 21% | 37% | 29% |

Percentages are reported within each expert group. Abbreviations: H = hearing loss experts; V = vision loss experts; H+V = cross-disciplinary experts.

Experts across both domains report a high volume of variants, with about half curating over 100 variants annually (**Fig. 1A**). Many variants are classified as VUS due to insufficient data and functional evidence resolves only a subset of these cases (**Fig. 1B-C**). Despite this, most respondents reclassify 1-10 variants each year, but only a subset is driven by new data (**Fig. 1D-E**). Conflicting functional evidence is a significant, common challenge, reported by 54% of participants (**Additional file 2: Fig. S1D**). Strategies for resolving conflicts include independent assessment of experimental design, adjusting the application of PS3/BS3 criteria, consulting peers, comparing with other evidence, performing additional experiments or following ClinGen guidance (**Table 2**). Taken together, it is suggested that functional evidence can aid variant classification, but barriers, including conflicting results and limited data, persist.

**Figure 1.**
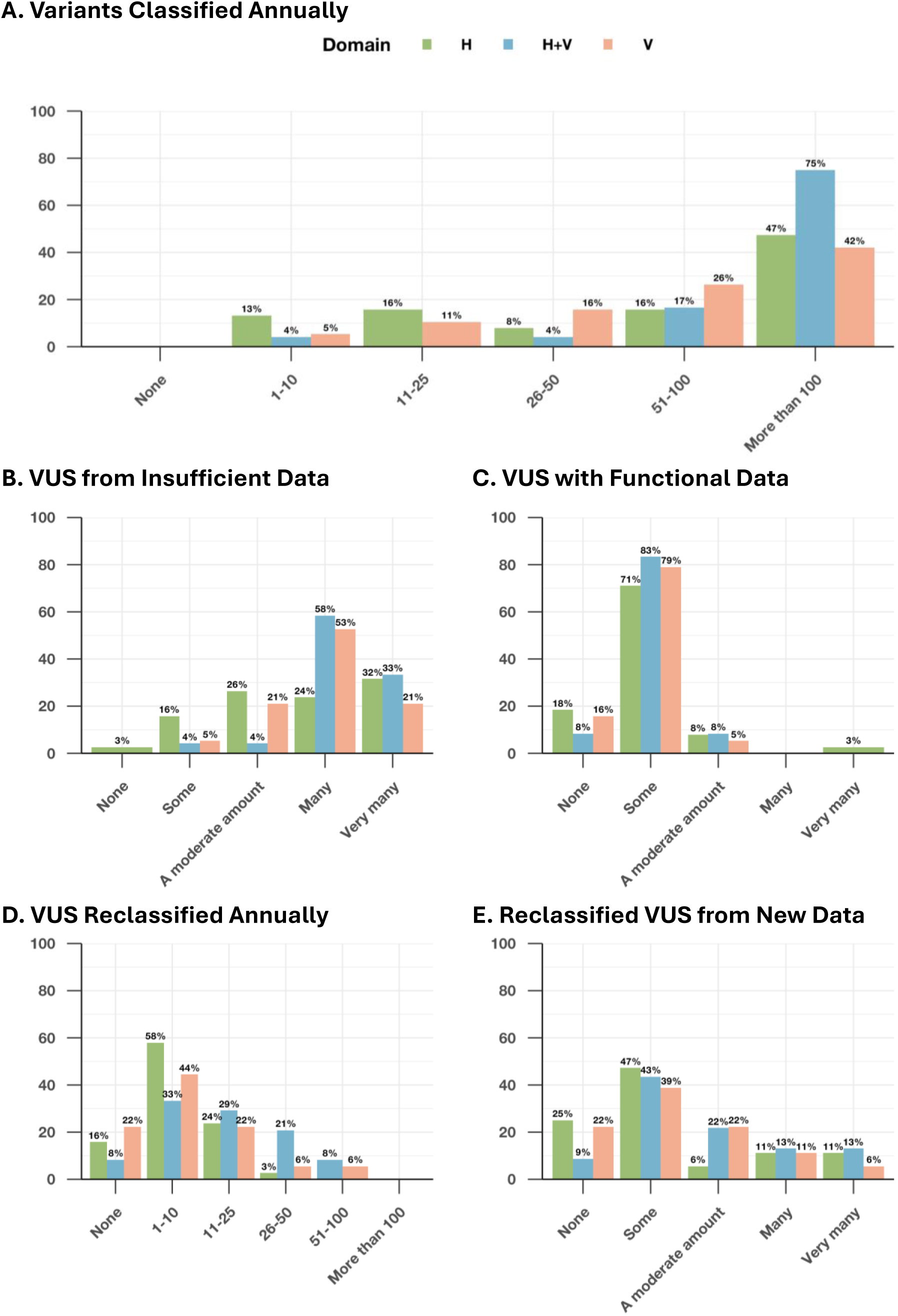
Scopes and challenges of variant interpretation by disease domain. Each subplot represents responses to a different survey question: (**A**) number of variants evaluated per year, (**B**) proportion of variants classified as VUS due to insufficient data, (**C**) proportion of VUS with available functional data used for classification, (**D**) number of VUS reclassified annually, (**E**) proportion of VUS reclassified specifically due to new or updated data. Bars are grouped by disease domain: H (hearing) shown in green, H+V (hearing and vision) in blue, and V (vision) in red.

**Table 2.** Approaches to handling conflicting functional evidence.

|  |  |
| --- | --- |
| Assess the experiment (*n* = 10) | "I am reviewing each functional experiment (material and methods) to conclude which is the strongest evidence."<br>"Assessment of the functional studies by considering the model organism(s) utilized, assay calibration, usage of negative and positive controls, type of experiments performed, impact factor of the publishing journal."<br>"I have weighed the corresponding papers, and made a calculated decision based on my own interpretation of the results (validity, specificity), taken together with the type of journal, the authors, etc." |
| Modify the guidelines (*n* = 7) | "Classified variant as VUS unless obvious problem with the data."<br>"Not applying either PS3 or BS3 at any level strength."<br>"Either don't use functional evidence at all, or modify PS3/BS3 applicable." |
| Discuss with peers (*n* = 4) | "The practice in ClinGen VCEPs is to consult the experts about which piece of functional evidence is more convincing, if any." |
| Compare with other evidence (*n* = 3) | "I integrate with other evidence and consider clinical, segregation, and computational data to support or refute functional findings." |
| Perform further experiments (*n* = 3) | "As I am lucky to work in a research genetic lab myself, I performed a new functional validation and confirmed one of the previous assessments." |
| Follow ClinGen specifications (*n* = 2) | "As part of a ClinGen VCEP, (...) if results from different assays are conflicting for a single variant, then the result from the assay with the highest level of validation and a conclusive result can override the result from the other assay." |

### Professional roles and collaborative engagement

We explored the professional roles of HL, VL, and combined H+V experts by asking about their participation in clinical and research activities. Most HL (69%) and H+V specialists (83%) perform variant classification in clinical settings, compared with 47% of VL experts, likely reflecting the higher proportion of research-focused respondents in the VL group (**Fig. 2A**). Research-based variant classification, discussing genetic test results with other labs and participating in multidisciplinary meetings were common across all groups (65-92%) (**Fig. 2B, 2E**). Notably, VL experts demonstrated a broader clinical communication role, more frequently reporting that they communicate results directly with patients (68-84% vs. 45-62%) (**Fig. 2C-D, 2F**). VL specialists also show higher current involvement in ClinGen Working Groups (74% vs. 25-37%) (**Fig. 2G**). These results suggest that HL, VL, and H+V specialists participate actively in research, variant classification, multidisciplinary discussions and collaborative efforts, underscoring their common commitment to improving the interpretation and application of genetic data.

**Figure 2.**
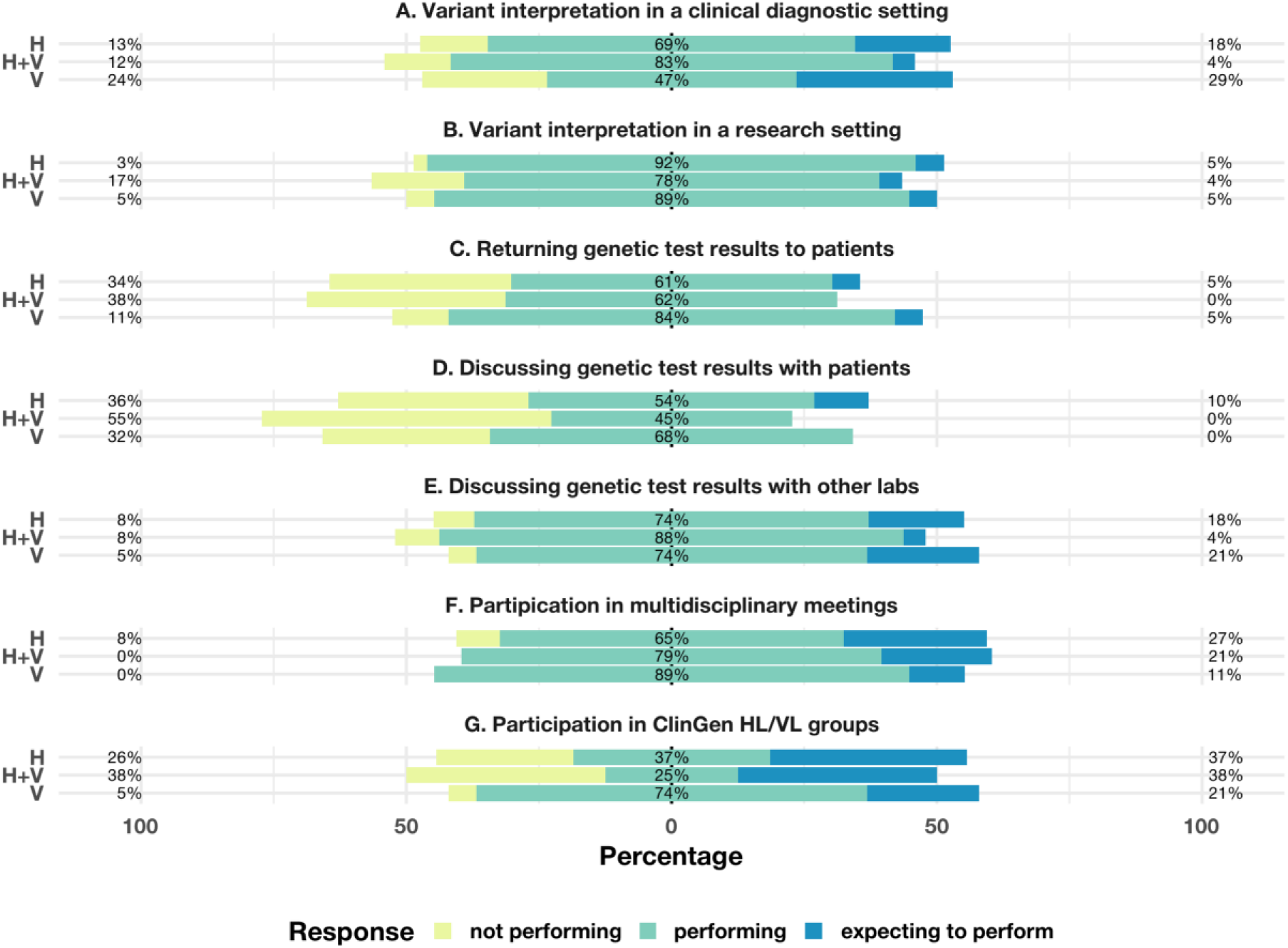
Current and anticipated professional activities. Survey respondents (H = hearing, V = vision, H+V = both hearing and vision) were asked about their current and expected participation in variant interpretation and related professional activities. Bars indicate the proportion of experts within each domain who reported not performing (yellow), currently performing (teal), or expecting to perform (blue) each activity (**A**-**G**).

### Engagement with functional evidence

Functional evidence can be broadly classified as diagnostic-grade, generated under clinically validated conditions, or research-grade, produced in research settings for exploratory purposes, but not necessarily meeting clinical validation standards. Professionals collaborate with other labs to request functional data that is not available from public sources or is not credible. To gain insight into how HL and VL professionals engage with functional evidence, we asked respondents which functional evidence-related tasks they are currently performing. Across all groups, most respondents reported using public data sources (58-87%), reassessing variants with functional evidence (68-83%) and curating functional data for variant classification (58-67%) (**Fig. 3A-C**). However, requests for diagnostic-grade functional evidence from clinical laboratories were relatively uncommon (37-45% vs. 43-74%), likely reflecting the limited availability of approved clinical assays for both HL- and VL-associated genes (**Fig. 3D-E**). Collaboration with research laboratories was common across domains, including those specializing in hearing or vision loss (57-63%) or in fundamental mechanisms and protein properties (38-51%) (**Fig. 3F-G**). Overall, HL and VL specialists actively engage with functional evidence through public resources and collaborations, with an emerging interest in expanding functional data use.

**Figure 3.**
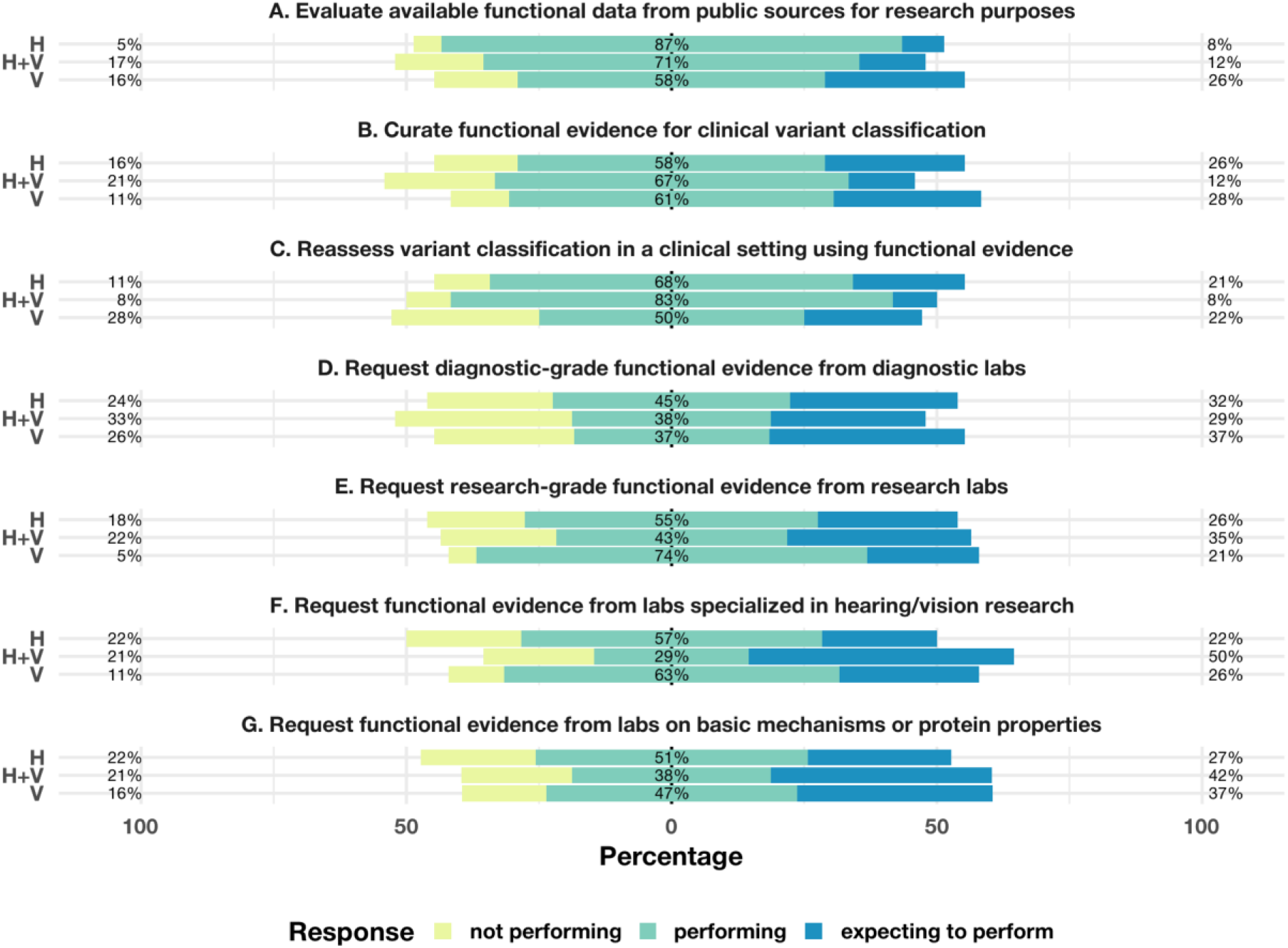
Current and anticipated functional evidence tasks. Survey respondents (n = 82) (H = hearing, V = vision, H+V = both hearing and vision) reported their involvement in tasks related to the use and generation of functional evidence. Bars indicate the proportion of experts within each domain who reported not performing (yellow), currently performing (teal), or expecting to perform (blue) each task (**A**-**G**).

### Confidence with functional evidence types and models

Respondents rated their confidence in applying different functional data types for variant classification, from *not confident at all* to *very confident*. Experts expressed moderate confidence in biochemical assays (47-63%) (**Fig. 4A**). Transcript-based assays, such as splicing assays, (53-75%) and patient-derived cell models (52-70%) were generally trusted across all respondents (**Fig. 4B-C**). Confidence in gene-edited *in vitro* models ranged from 46-69% (**Fig. 4D**). Complex *in vivo* systems, including various animal models, were trusted by 37-67% of respondents (**Fig. 4F-H**), while less complex models, such as cell models for genes of uncertain significance, zebrafish, and *Drosophila* models, were met with moderate to low confidence across all groups (11-54%) (**Fig. 4E, 4I-M**). These results indicate that respondents had highest confidence in biochemical and transcript-based assays, moderate confidence in gene-edited *in vitro* models and more variable confidence in complex *in vivo* systems.

**Figure 4.**
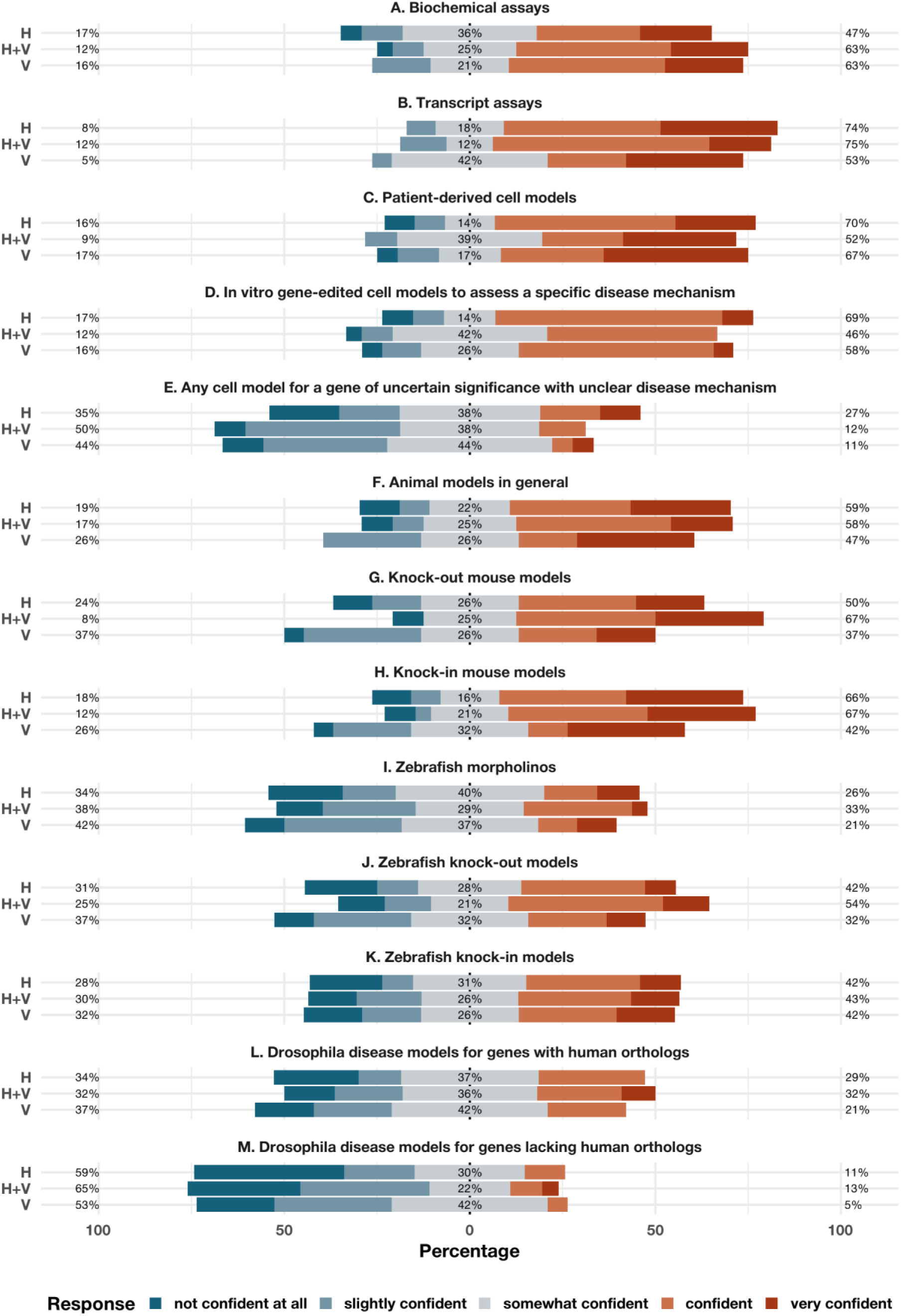
Confidence in using functional evidence. Stacked bar plots display confidence levels reported in applying various types of functional assays for variant interpretation (**A**-**M**) across disease domains: hearing loss (H), ocular diseases (V), and both domains (H+V). Responses were collected on a five-point Likert scale and grouped into three percentage categories: (i) not confident/slightly confident (blue), (ii) somewhat confident (grey), and (iii) confident/very confident (red). Missing responses (1-6 per item) were not shown.

### Familiarity and confidence in functional evidence resources

Experts rated their confidence in utilizing a variety of available functional evidence resources. The highest confidence was consistently reported for functional data found in the primary literature and established literature-based clinical databases such as ClinVar [9], LitVar2 [23], and OMIM [24] (39-74%) (**Fig. 5A-B**). Mouse model databases were perceived with moderate confidence (26-49%) (**Fig. 5C**), non-mammalian resources (e.g., ZFIN [25], FlyBase [26], Xenbase [27]) were generally unfamiliar or unreliable (9-32%) (**Fig. 5D-G**). Computational predictors such as SpliceAI [28], AlphaMissense [29], and REVEL [30] were viewed with moderate to high confidence (42-67%) (**Fig. 5H**). Trust in MaveDB [16] was limited across all domains (11-26%) (**Fig. 5I**). Broadly, these insights suggest that experts have highest confidence in published and literature-curated sources for functional data and computational predictors, while confidence in non-mammalian functional data and high-throughput experimental assay data was comparatively limited.

**Figure 5.**
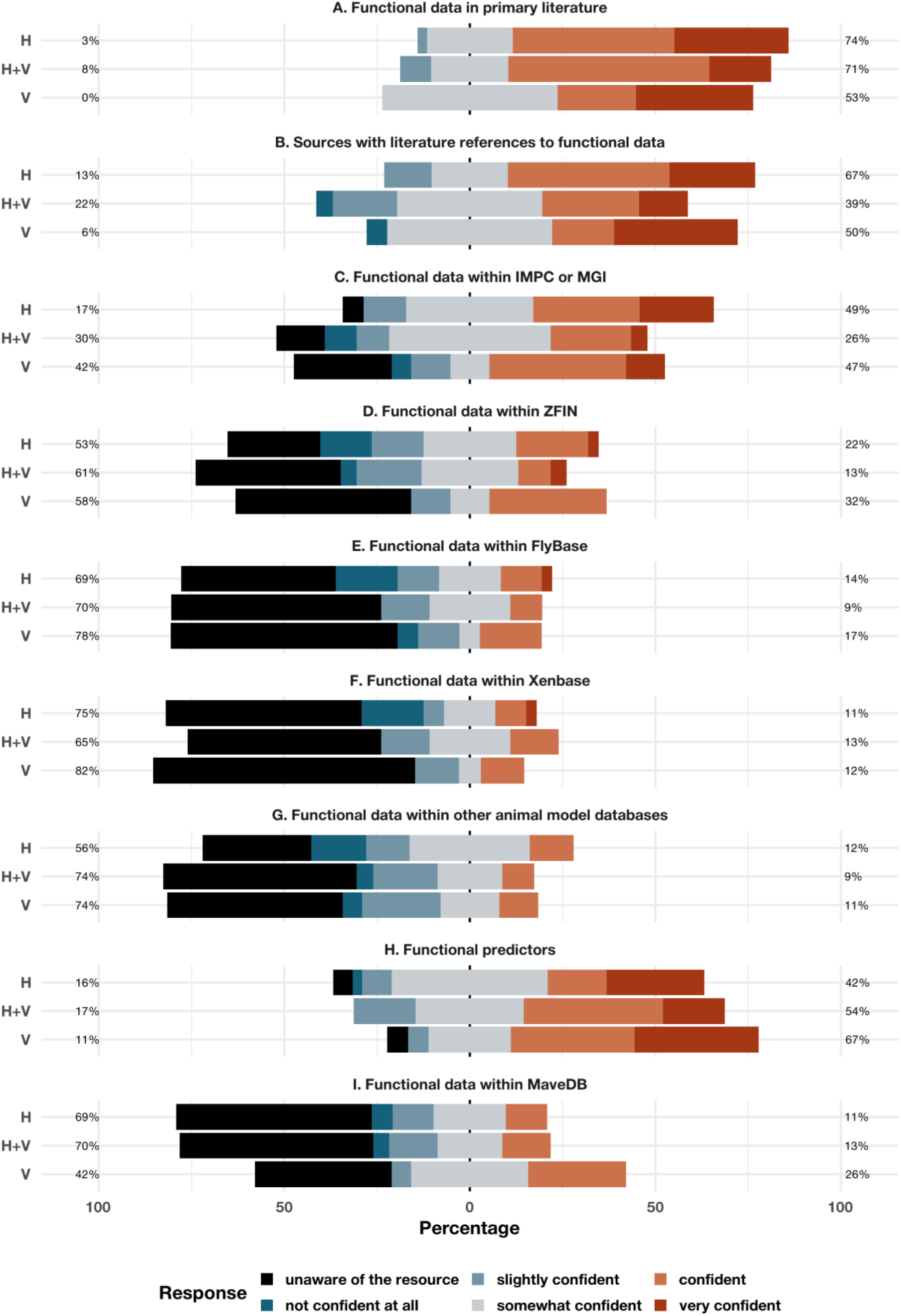
Confidence in functional evidence resources. Stacked bar plots display confidence levels reported in functional evidence resources (**A**-**I**) across disease domains: hearing loss (H), ocular diseases (V), and both domains (H+V). Responses were collected on a six-point Likert scale and grouped into three percentage categories: (i) not confident/slightly confident (black/blue), (ii) somewhat confident (grey), and (iii) confident/very confident (red). Missing responses (1-6 per item) were not shown.

### Confidence in standardized criteria

Confidence in applying PS3/BS3 criteria was assessed by their comfort level with relevant guidelines. Relatively high confidence (50-89%) was reported across the board for the ACMG/AMP Guidelines, ClinGen PS3/BS3 specifications, ClinGen splicing evidence recommendations and the ClinGen functional assessment worksheet (**Fig. 6A-C, 6E**). Both HL and VL experts found domain-specific guidelines moderately useful (38-68%) (**Fig. 6D, 6F-I**). However, confidence in the ClinGen glaucoma guidelines was lower among VL specialists (37%), with some unaware of these recommendations (**Fig. 6J**). This discrepancy likely reflects the relative timing of guideline development, as the ocular domain was established more recently. In open-ended responses, one participant reported using X-linked inherited retinal disease specifications for *RPGR* and *RS1* in addition to the listed guidelines in this survey. Generally, specialists in HL and VL show broad familiarity with functional evidence guidelines.

**Figure 6.**
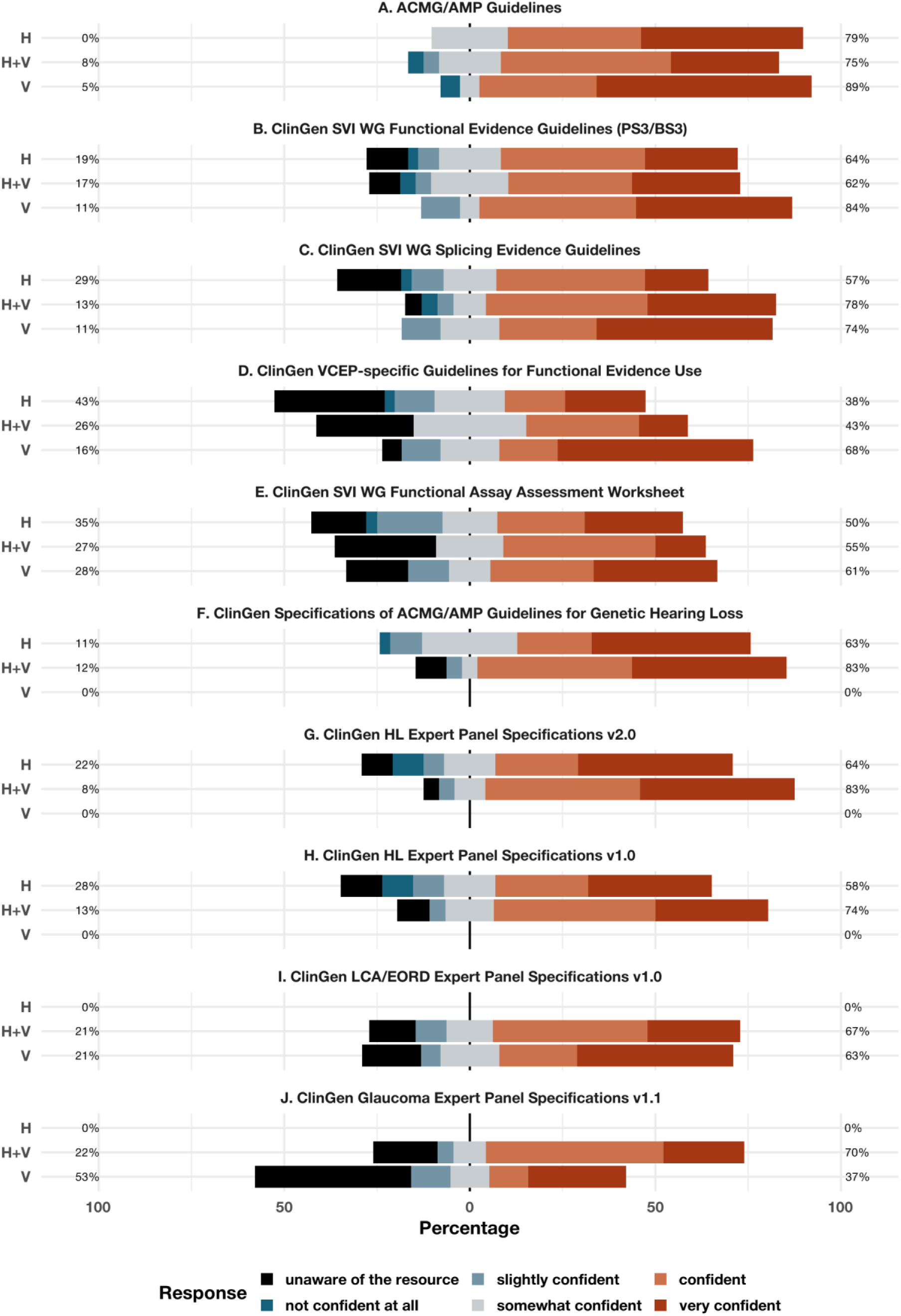
Confidence in variant interpretation guidelines. Stacked bar plots display confidence levels reported in interpretation guidelines (**A**-**J**) across disease domains: hearing loss (H), ocular diseases (V), and both domains (H+V). Responses were collected on a six-point Likert scale and grouped into three percentage categories: (i) not confident/slightly confident (black/blue), (ii) somewhat confident (grey), and (iii) confident/very confident (red). Missing responses (1-8 per item) were not shown.

### Improving and assessing the functional evidence application

**Barriers to functional evidence application:** Respondents rated the extent to which various barriers impact their use of functional data. Insufficient training, both general and disease-specific, was reported as an occasional challenge (21-42%) (**Additional file 2: Fig. S2A-S2E**). The most frequently reported challenge among HL experts was interpreting reduced penetrance or variable expressivity (58%) (**Additional file 2: Fig. S2F**). Concerns regarding the quality and accuracy of functional evidence were frequently reported (50-68%) (**Additional file 2: Fig. S2G-S2H**), as were challenges in locating and interpreting functional data in literature and variant databases (17-53%) (**Additional file 2: Fig. S2I-S2L**). Understanding clinically relevant transcripts for HL and VL-related genes also presented a moderate challenge (31-42%) (**Additional file 2: Fig. S2M**). Additional challenges cited by respondents in an open-ended section included the substantial time and effort required to evaluate assay validity and relevance, variability in guideline application, limited genetics knowledge among clinicians, and restricted access to publications.

**Strategies for improvement:** To understand how the use of functional evidence could be enhanced, respondents rated the impact of various factors. Workshops, additional VCEP specifications and disease-specific guidelines were widely regarded as useful (54-83%) (**Additional file 2: Fig. S3A, S3E-S3G**). Online training modules and training spreadsheets were rated as helpful (42-69%) (**Additional file 2: Fig. S3B-S3D**). Improved access to primary functional data and standardized assessments through ClinVar was viewed as highly beneficial (72-96%) (**Additional file 2: Fig. S3H-S3I**). Furthermore, the ability to request the *de novo* generation of functional data for specific variants or access gene-wide deep mutational scans was consistently rated as impactful for enhancing variant classification (58-86%) (**Additional file 2: Fig. S3J-S3K**).

### Expectations about clinical utility and future data use

**Clinical utility and data curation needs:** Respondents indicated their level of agreement with several statements regarding the role of functional evidence in clinical interpretation. Opinions were mixed on whether existing literature provides sufficient support for the clinical utility of variant-level functional data or MAVE datasets (**Fig. 7A-B**). Across all groups, there was a consensus on the need for additional studies demonstrating clinical utility and for clarifying clinically relevant transcripts in ClinVar (53-100%) (**Fig. 7C-E**).

**Figure 7.**
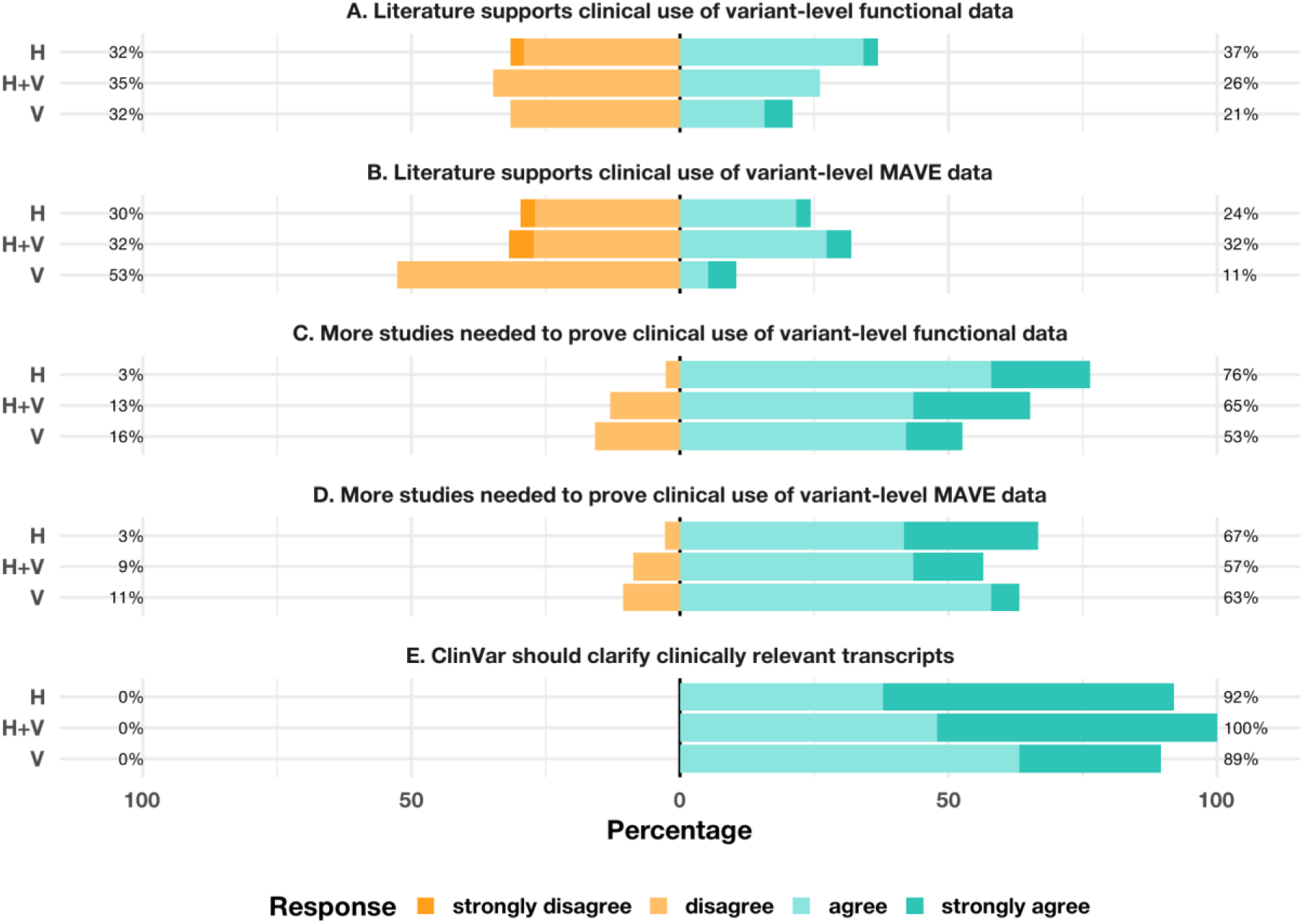
Assessment of existing data on improving clinical utility. Stacked bar plots display agreement with statements regarding interaction with functional assays to improve clinical utility (**A**-**E**) across disease domains: hearing loss (H), ocular diseases (V), and both domains (H+V). Responses were collected on a five-point Likert scale, ranging from strongly disagree (orange) to strongly agree (cyan), and grouped into two percentage categories: (i) strongly disagree/disagree, and (ii) agree/strongly agree. Missing responses (2-4 per item) were not shown. Neutral responses are excluded from display.

**Future expectations for functional data integration:** HL and VL experts were asked about their current practices and future expectations regarding functional data use. They reported reviewing literature for both gene- and variant-level functional evidence during variant classification (78-91%) (**Additional file 2: Fig. S4A, S5A**). Gene-level data was considered highly helpful (74-87%) in the absence of variant-specific data (**Additional file 2: Fig. S4B-S4C**). Experts identified essential requirements for the proper use of variant-level functional data in ClinVar, including the specification of detailed methods as well as disease mechanism (65-95%) (**Additional file 2: Fig. S5B-S5C**). Metrics reflecting assay credibility (e.g., reproducibility, assay type, quality control measures), linked directly to primary data sources and by showing multiple resources, even if they conflict, were seen as beneficial (61-100%) (**Additional file 2: Fig. S5D-S5G**). A majority stated they would adopt pre-defined functional evidence levels (e.g., PS3_Strong or PS3_Supporting) if integrated into ClinVar (53-85%) (**Additional file 2: Fig. S5H**).

## Discussion

We captured the perspectives of 82 professionals on the use of functional evidence for variant classification for HL and VL, representing the first survey focused on non-cancer hereditary disorders. Although functional evidence is expected to play a central role in resolving VUS, challenges remain in accessing high-quality functional data, interpreting complex methodological details, and integrating these findings into clinical decisions. Our findings emphasize the ongoing need for improved data transparency and structured evidence curation to strengthen variant classification practices.

HL and VL specialists demonstrated contrasting comfort level with different types of functional assays. HL specialists were generally more hesitant to use biochemical assays for variant classification; this is likely reflecting a perceived lack of reliable and physiologically relevant assays systems for many HL-associated genes. This may stem from the limited availability of physiologically relevant assay systems and the stringent specifications adopted by HL Clinical Domain Working Group (CDWG). The ClinGen HL CDWG reserves PS3_Strong for knock-in mouse models, only specific biochemical assay types would achieve PS3 evidence. For example, for *GJB2*, electrical coupling and dye transfer assays were considered to provide moderate-level evidence [31]. On the other hand, the Ocular CDWG approved a broader range of *in vitro* functional assays as strong-level evidence for genes implicated in glaucoma, Leber Congenital Amaurosis/early onset Retinal Dystrophy [32] and X-linked Inherited Retinal Disorders [33]. Given the anatomical and physiological differences between rodent and human ocular structures [34], it is not surprising that VL experts put a greater emphasis on *in vitro* assays and computational predictors than *in vivo* models. Furthermore, HL and VL experts jointly expressed less trust in large-scale functional studies and corresponding functional evidence frameworks. Such tendency may reflect the limited knowledge of the molecular mechanisms of many HL- and VL-associated genes, as functional assays might not account for the full biological complexity (e.g., hair cell biology) that may be impacted. Taken together, these findings indicate the need for disease-specific functional evidence frameworks that can better accommodate the distinct biological and methodological challenges of each domain.

HL experts specifically reported challenges related to the application of functional evidence when the possibility of reduced penetrance or variable expressivity exists. Several cases of incomplete penetrance and modifier effects have been previously reported for HL [35–38]. While large-scale functional studies like MAVEs are expected to help resolve many VUSs, they are not anticipated to resolve all VUS due to biological heterogeneity and mechanistic complexity caused by variable penetrance [39]. To address these limitations, groups such as the Cancer Variant Interpretation Group UK (CanVig-UK) have proposed frameworks to interpret variants of reduced penetrance, particularly those manifesting intermediate or conflicting phenotypes in functional assays [40]. Given that gene regulatory variation can modify penetrance [41], it is also important to integrate insights from high-throughput assays for regulatory variation, such as massively parallel reporter assays (MPRAs) [42]. Moreover, emerging guidelines specifically for interpreting non-coding variants may provide valuable insights into how these variants influence gene expression and disease risk [43]. These multifaceted approaches may offer a more comprehensive strategy for reducing uncertainty in variant classification by complementing findings from large-scale functional assays.

Considering that the term VUS encompasses variants with a broad range of posterior probabilities of pathogenicity (10-90%), subclassifying them could further refine clinical decision-making processes. Supporting this approach, a framework for VUS subclassification integrated by four clinical laboratories demonstrated the potential to substantially reduce uncertainty, with published data indicating that VUS sub-tiering resolved approximately 40% (23,067/57,480) of VUS in a large cohort [44]. Furthermore, it has been previously shown that HL- and inherited retinal disorder (IRD)-specific guidelines improved variant classification, resolving 62% [45] and 53% [46] of VUS, respectively. These results strongly suggest that the revision of existing guidelines and integration of them with domain-specific recommendations can dramatically reduce uncertainty in variant classification.

The earlier survey of 190 genetics professionals [21] and our survey of 82 domain experts in hearing and vision loss revealed several consistent themes. Both studies emphasize that limited availability, conflicting results and uncertainty about assay validity are major barriers to incorporating functional data into variant interpretation. A pervasive lack of confidence in the quality and accuracy of available assays was identified as a major barrier, depicting the importance of resources like MaveMD [47], which integrates MAVE datasets with clinical evidence and structured outputs compatible with the ACMG/AMP guidelines. Participants in both studies expressed strong support for improved standardization, harmonized frameworks, and better access to primary functional data, as well as for integrating standardized PS3/BS3 interpretations into widely used resources such as ClinVar, to enhance the reliability and consistency of variant classification.

## Conclusions

Our findings underline that both shared and disease-specific challenges exist in leveraging functional evidence for variant classification in HL and VL. While experts across both domains recognize the value of functional data, they identify barriers related to assay quality, lack of standardized assay systems and evidence evaluation criteria and limited data accessibility. Addressing these gaps through improved data sharing, methodological transparency and specialized training will be essential to fully realize the clinical utility of functional evidence in variant classification.

## Data Availability

The survey data and scripts used to generate the figures in this study are available through GitHub: https://github.com/ardainn/VUS-Hearing-Ocular/

## Abbreviations

ACMG: American College of Medical Genetics and Genomics
AMP: Association of Molecular Pathology
BS3: Benign strong 3
CanVig-UK: Cancer Variant Interpretation Group UK
CDWG: Clinical Domain Working Group
ClinGen: Clinical Genome Resource
GCEP: Gene Curation Expert Panels
H+V: Combined hearing loss and vision loss
HL: Hearing loss
IMPC: International Mouse Phenotyping Consortium
IRD: inherited retinal disorder
LCA/EORD: Leber Congenital Amaurosis/early onset Retinal Dystrophy
MAVE: Multiplexed assays of variant effects
MGI: Mouse Genome Informatics
MPRA: Massively parallel reporter assay
OMIM: Online Mendelian Inheritance in Man
PS3: Pathogenic strong 3
REDCap: Research Electronic Data Capture
REVEL: Rare Exome Variant Ensemble Learner
RNA: Ribonucleic acid
SVI: Sequence Variant Interpretation
VCEP: Variant Curation Expert Panels
VL: Vision loss
VUS: Variant of uncertain significance
WG: Working group
ZFIN: Zebrafish Information Network

## Data Availability

The survey data and scripts used to generate the figures in this study are available through Github: https://github.com/ardainn/VUS-Hearing-Ocular/

## Acknowledgements

We thank the participants for taking the time to complete our survey. This work was done with the support of the Center for Rare Hearing Disorders at the Center of Rare Diseases Göttingen (ZSEG). We thank Bernd Wollnik for early feedback.

## Author information

<u>Contributions</u>: Analysis and writing of the first draft: RAI and BV. Study design and implementation: AAT, SSA, MD, RBH, and BV. TB, LMS, and ABS supported interpretation. All authors have edited and approved the final draft.

Corresponding author: Correspondence to Barbara Vona.

## Ethics declarations

Ethics approval: The ethics committee of the University Medical Center Göttingen gave ethical approval for this work (approval number 12/8/24 An).

## Supplementary information

Additional file 1: Survey questionnaire used in the study (.pdf).

<u>Additional file 2</u>: Supporting Figures S1-S5 (.docx). Fig S1: Participant characteristics and experiences with functional evidence. Fig S2: Challenges in using functional evidence for variant interpretation. Fig S3: Approaches to improving the interaction and application of functional evidence. Fig S4: Use and expectations of gene-level functional data. Fig S5: Use and expectations of variant-level functional data.

## Additional File 1

### Assessment of Current and Future Approaches to Address Variants of Uncertain Significance in Hearing and Ocular Genomics Domains

Purpose of the QuestionnaireThis questionnaire aims to explore the practices and experiences of professionals in the hearing loss and ocular genetics domains who are actively incorporating variant-level functional data into workflows. By focusing on these areas, we seek to gather diverse perspectives on variants of uncertain significance (VUS) and better understand how current and future functional data will be integrated into clinical practice.We aim to address key topics, including: - The impact of VUS in routine workflows - The practical applications and challenges of using functional evidence criteria in variant classification - The level of confidence in applying functional evidence criteria - Expectations for the type of information included when aggregating variant classification evidence at a single source like ClinVar - Perspectives for future inclusion of high-throughput approaches, such as multiplexed assays of variant effect (MAVEs), into resources like ClinVar Our goal is to enhance understanding of the current awareness and application of functional data for interpreting VUS within these two clinical domains. We join international efforts to address the challenges posed by VUS. Functional data have conventionally been generated on a variant-by-variant basis using cell or animal models. However, scalable functional assays like MAVEs offer the potential to accelerate VUS reclassification, providing more certainty about whether variants are pathogenic or benign. This study seeks to capture current and future perspectives on these developments from active members of the hearing loss and ocular genetics communities. Estimated time for completion:15-20 minutes Ethics Approval:This study was approved by the Ethics Committee of the University Medical Center Göttingen (Approval: 12/8/24, Study Center ID:

2024-03475). Acknowledgement:We acknowledge Dr. Lea M. Starita and Dr. Andrew B. Stergachis from the Department of Genome Sciences, University of Washington, Seattle, WA, USA for inspiring this work with their preprint (https://doi.org/10.1101/2025.01.25.25321117). We appreciate your participation and willingness to contribute to this important work. Sincerely,Dr. Ahmad Abou TayounAl Jalila Genomics Center of Excellence, Al Jalila Children’s Specialty Hospital, Dubai Health, Dubai, UAEDr. Sami S. AmrLaboratory for Molecular Medicine, Mass General Brigham Personalized Medicine, Cambridge, Massachusetts, USA Department of Pathology, Brigham and Women’s Hospital, Boston, Massachusetts, USADeparment of Pathology, Harvard Medical School, Boston, Massachusetts, USADr. Marina DiStefanoMedical and Population Genetics, Broad Institute of MIT and Harvard, Cambridge, Massachusetts, USADr. Robert B. HufnagelCenter for Integrated Healthcare Research, Kaiser Permanente, Hawai’i Region, Honolulu, HI, USA Ophthalmic Genetics and Visual Function Branch, National Eye Institute, National Institutes of Health, Bethesda, MD, USADr. Barbara VonaInstitute for Auditory Neuroscience, University Medical Center Göttingen, Göttingen, Germany

**Consent, Participation Eligibility, Data Use, and Contact Information**

Consent We are conducting a study on the impact of variants of uncertain significance (VUS) and variant-level functional data. We are seeking insights and experiences from active hearing and ocular genetics professionals whose expertise are invaluable to our comprehensive understanding of the challenges and expectations of these communities.

Eligibility for participation We request that only individuals actively involved in clinical variant classification in hearing loss and/or retinal diseases participate in this questionnaire.

Participation is voluntary and anonymity is guaranteed

Participation is voluntary and your responses are anonymous. If you decide not to continue while completing the questionnaire, you can exit the page at any time to discontinue participation. By submitting the questionnaire, you consent to the use of your data. Please note that once submitted, responses cannot be deleted due to the anonymous nature of data collection. Use of questionnaire data The anonymous data collected through this questionnaire will be used to explore current and future challenges faced by the hearing and ocular genetics communities. These data will be analyzed and contribute to the development of educational resources. Summarized responses may be published in peer-reviewed journals, included in educational materials, or be presented at conferences. All reponses are anonymous. We do not collect information that would identify participants. The questionnaire is estimated to take 15-20 minutes of your time. While participant perspective are vital in guiding the use of functional data in hearing and ocular genomic medicine and future development of databases, there is no immediate direct benefit for participating. However, the goal is that your participation will help improve the accessibility and utility of genomic databases, ultimately advancing scientific progress in hearing and ocular genomic medicine.

Contact information Please contact the study leader (Dr. Barbara Vona) in case of questions or concerns.

Thank you for your participation!

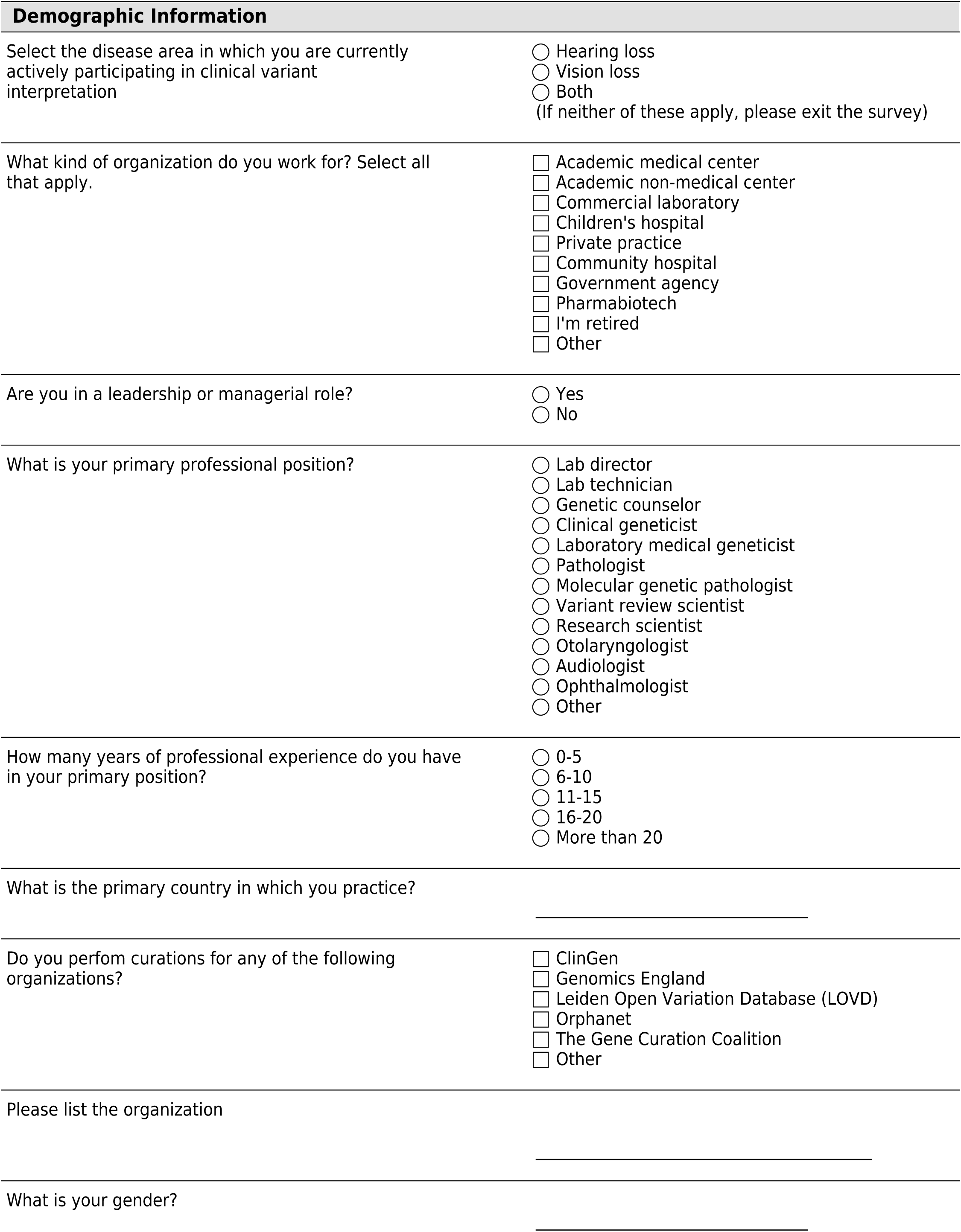

|  | I am not performing and will not perform in future | I am currently performing | I am not performing now but expect to in the future |
| --- | --- | --- | --- |
| Variant interpretation in a clinical diagnostic setting | ☐ | ☐ | ☐ |
| Variant interpretation in a research setting | ☐ | ☐ | ☐ |
| Returning genetic test results to patients | ☐ | ☐ | ☐ |
| Discussing genetic test results with patients | ☐ | ☐ | ☐ |
| Discussing genetic test results with other labs | ☐ | ☐ | ☐ |
| Participation in multidisciplinary meetings (e.g. hearing or vision genetics meetings) | ☐ | ☐ | ☐ |
| Participation in the ClinGen Hearing Loss Expert Panel or the ClinGen Ocular Clinical Domain Working Group, including any of the Variant Curation Expert Panels (VCEPs) or Gene Curation Expert Panels (GCEPs) | ☐ | ☐ | ☐ |

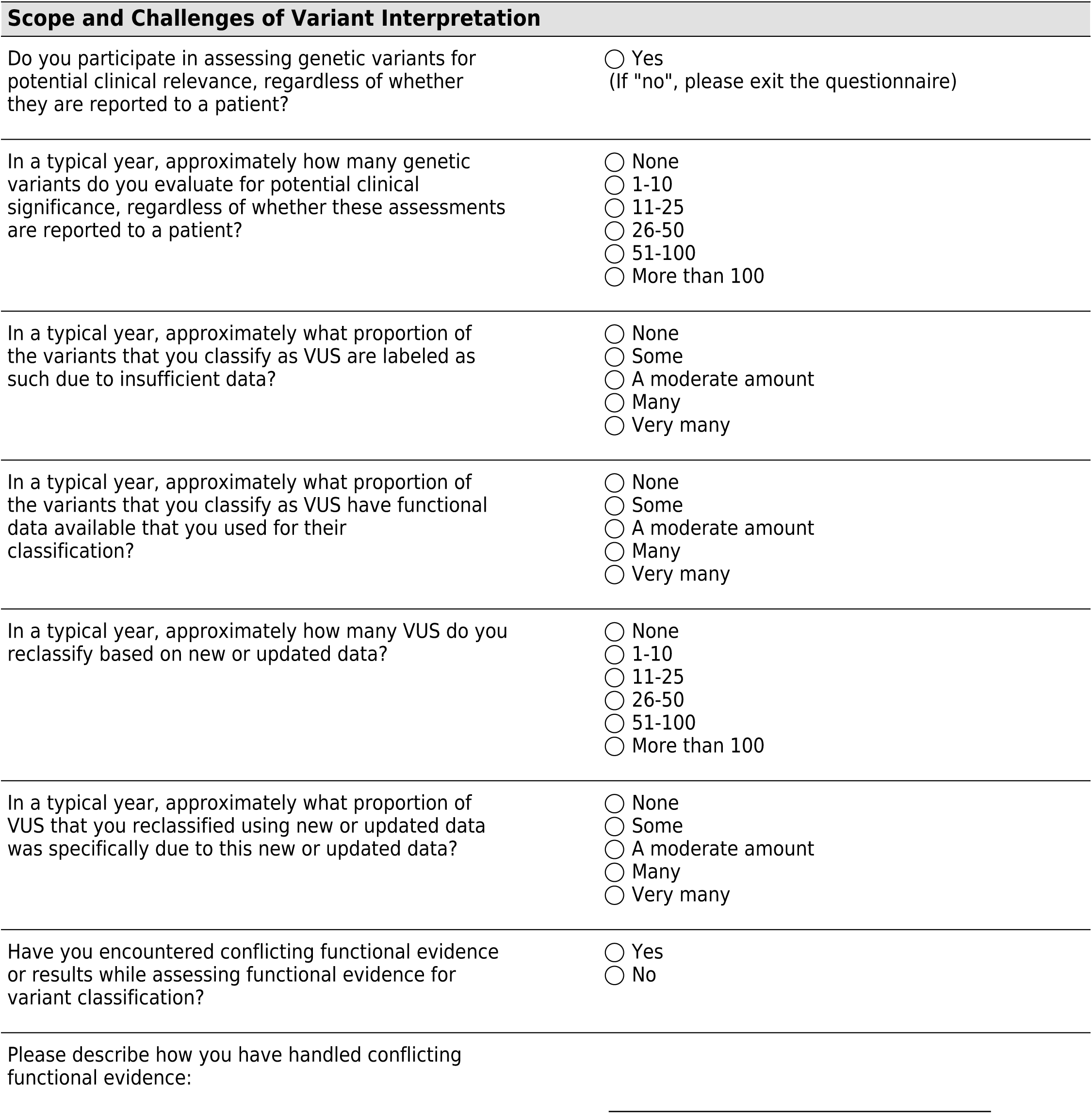

|  | I am not performing and will not perform in future | I am currently performing | I am not performing now but expect to in the future |
| --- | --- | --- | --- |
| Evaluate available functional data from public sources for research purposes | ☐ | ☐ | ☐ |
| Curate functional evidence for clinical variant classification | ☐ | ☐ | ☐ |
| Reassess variant classification in a clinical setting using functional evidence | ☐ | ☐ | ☐ |
| Request diagnostic-grade functional evidence from diagnostic labs | ☐ | ☐ | ☐ |
| Request research-grade functional evidence from research labs | ☐ | ☐ | ☐ |
| Request functional evidence from research labs specifically specialized in hearing and/or vision functional research, irrespective of diagnostic- or research-grade | ☐ | ☐ | ☐ |
| Request functional evidence from research labs specialized in the basic mechanism or protein structural or biophysical properties, not necessarily from the hearing or vision fields, irrespective of diagnostic- or research-grade | ☐ | ☐ | ☐ |

Are there other activities involving functional evidence not listed above that you would like to add?

|  | 1 (Not confident at all) | 2 | 3 (Somewhat confident) | 4 | 5 (Very confident) |
| --- | --- | --- | --- | --- | --- |
| Biochemical assays (e.g. enzymatic activity assays) | ☐ | ☐ | ☐ | ☐ | ☐ |
| Transcript assays (e.g. splicing assays or transcriptomics data) | ☐ | ☐ | ☐ | ☐ | ☐ |
| Patient-derived cell models | ☐ | ☐ | ☐ | ☐ | ☐ |
| In vitro gene edited cell models to assess a specific disease mechanism (e.g. loss-of-function) | ☐ | ☐ | ☐ | ☐ | ☐ |
| Any cell model for a gene of uncertain significance with unclear disease mechanism | ☐ | ☐ | ☐ | ☐ | ☐ |
| Animal models in general (e.g. mouse or zebrafish models of genetic diseases) | ☐ | ☐ | ☐ | ☐ | ☐ |
| Knock-out mouse models | ☐ | ☐ | ☐ | ☐ | ☐ |
| Knock-in mouse models | ☐ | ☐ | ☐ | ☐ | ☐ |
| Zebrafish morpholinos | ☐ | ☐ | ☐ | ☐ | ☐ |
| Zebrafish knock-out models | ☐ | ☐ | ☐ | ☐ | ☐ |
| Zebrafish knock-in models | ☐ | ☐ | ☐ | ☐ | ☐ |
| Drosophila disease models for genes with human orthologs | ☐ | ☐ | ☐ | ☐ | ☐ |
| Drosophila disease models for genes lacking human orthologs | ☐ | ☐ | ☐ | ☐ | ☐ |

Are there additional types of functional evidence beyond those mentioned above that you utilize in genetic variant classification? Please specify particular assay categories and explain their importance in providing functional data.

|  | NA - I am<br>unaware of<br>the resource | 1 (Not<br>confident at<br>all) | 2 | 3 (Somewhat<br>confident) | 4 | 5 (Very<br>confident) |
| --- | --- | --- | --- | --- | --- | --- |
| Functional data in primary literature | ☐ | ☐ | ☐ | ☐ | ☐ | ☐ |
| Sources with literature references to functional data (e.g. ClinVar, LitVar2, OMIM) | ☐ | ☐ | ☐ | ☐ | ☐ | ☐ |
| Functional data within the International Mouse Phenotyping Consortium (IMPC) or Mouse Genome Informatics (MGI) | ☐ | ☐ | ☐ | ☐ | ☐ | ☐ |
| Functional data within the Zebrafish Information Network (ZFIN) database | ☐ | ☐ | ☐ | ☐ | ☐ | ☐ |
| Functional data within FlyBase | ☐ | ☐ | ☐ | ☐ | ☐ | ☐ |
| Functional data within Xenbase | ☐ | ☐ | ☐ | ☐ | ☐ | ☐ |
| Functional data within other animal model databases | ☐ | ☐ | ☐ | ☐ | ☐ | ☐ |
| Functional predictors such as SpliceAI, AlphaMissense, and/or REVEL | ☐ | ☐ | ☐ | ☐ | ☐ | ☐ |
| Functional data within MaveDB | ☐ | ☐ | ☐ | ☐ | ☐ | ☐ |

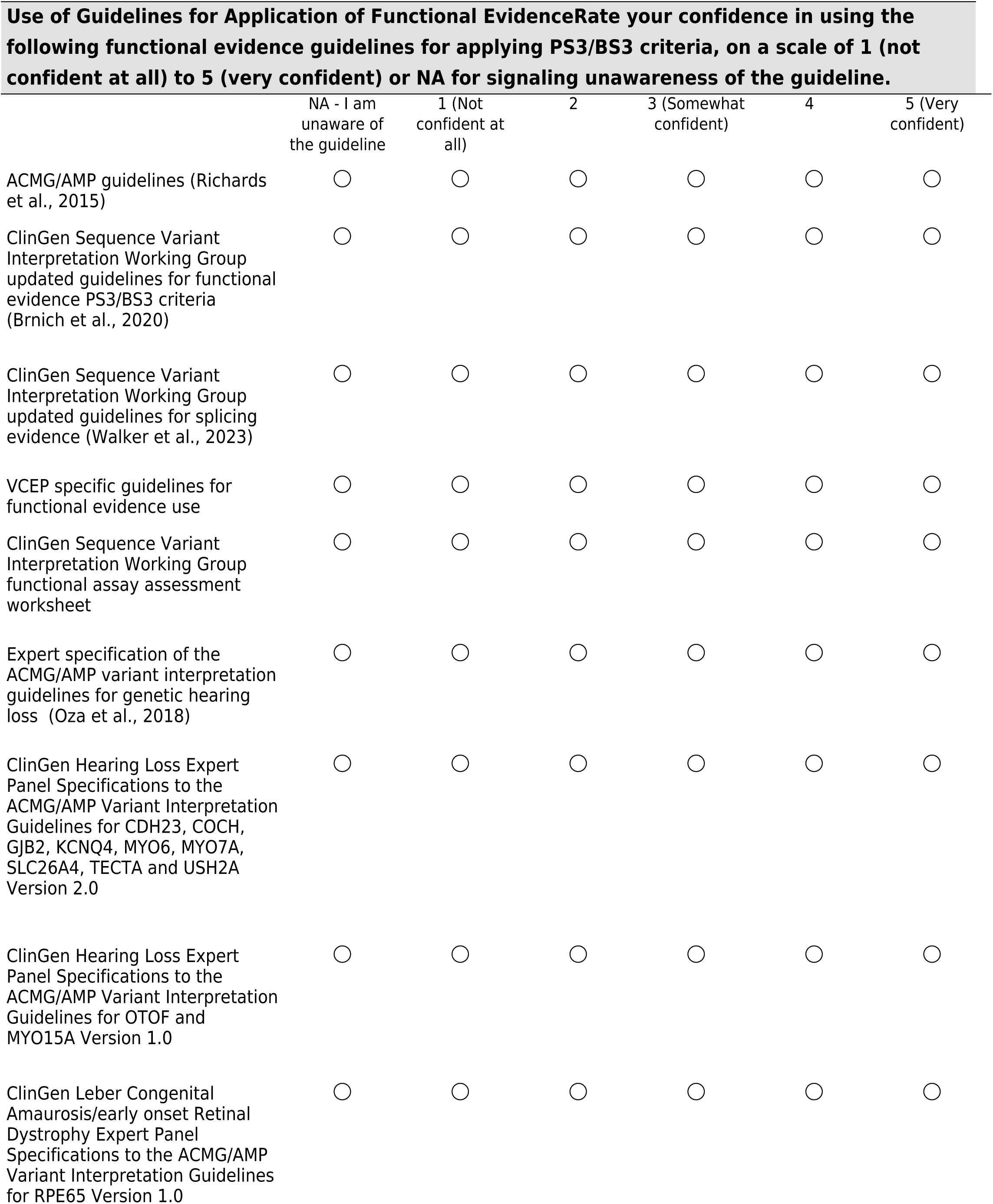
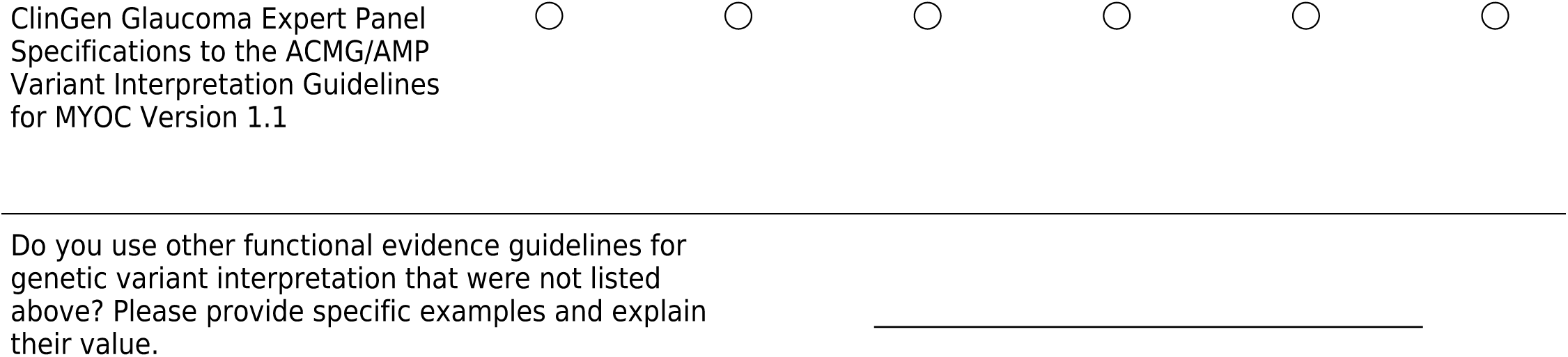

|  | 1 (Not a challenge for using functional evidence) | 2 | 3 (Occasionally a challenge for using functional evidence) | 4 | 5 (A significant challenge for using functional evidence) |
| --- | --- | --- | --- | --- | --- |
| Insufficient training on the general use of functional evidence for variant classification | ☐ | ☐ | ☐ | ☐ | ☐ |
| Insufficient training on the use of functional evidence for variant classification of loss-of-function variants | ☐ | ☐ | ☐ | ☐ | ☐ |
| Insufficient training on the use of functional evidence for variant classification of gain-of-function variants | ☐ | ☐ | ☐ | ☐ | ☐ |
| Insufficient training on the use of functional evidence for variant classification of dominant negative variants | ☐ | ☐ | ☐ | ☐ | ☐ |
| Insufficient training on the use of functional evidence for variant classification where haploinsufficiency is a possibility | ☐ | ☐ | ☐ | ☐ | ☐ |
| Insufficient training on the use of functional evidence for variant classification where reduced penetrance or variable expressivity is a possibility | ☐ | ☐ | ☐ | ☐ | ☐ |
| Insufficient information on the quality of functional evidence for variant classification | ☐ | ☐ | ☐ | ☐ | ☐ |
| Insufficient confidence in the accuracy of functional evidence for variant classification | ☐ | ☐ | ☐ | ☐ | ☐ |
| Challenges with locating functional evidence in primary literature | ☐ | ☐ | ☐ | ☐ | ☐ |
| Challenges with locating functional evidence in variant databases | ☐ | ☐ | ☐ | ☐ | ☐ |
| Challenges with understanding functional evidence found in primary literature | ☐ | ☐ | ☐ | ☐ | ☐ |
| Challenges with understanding functional evidence found in variant databases | ☐ | ☐ | ☐ | ☐ | ☐ |
| Challenges with understanding clinically relevant transcripts for hearing and/or vision | ☐ | ☐ | ☐ | ☐ | ☐ |

Please expand on any other significant challenges not described above.

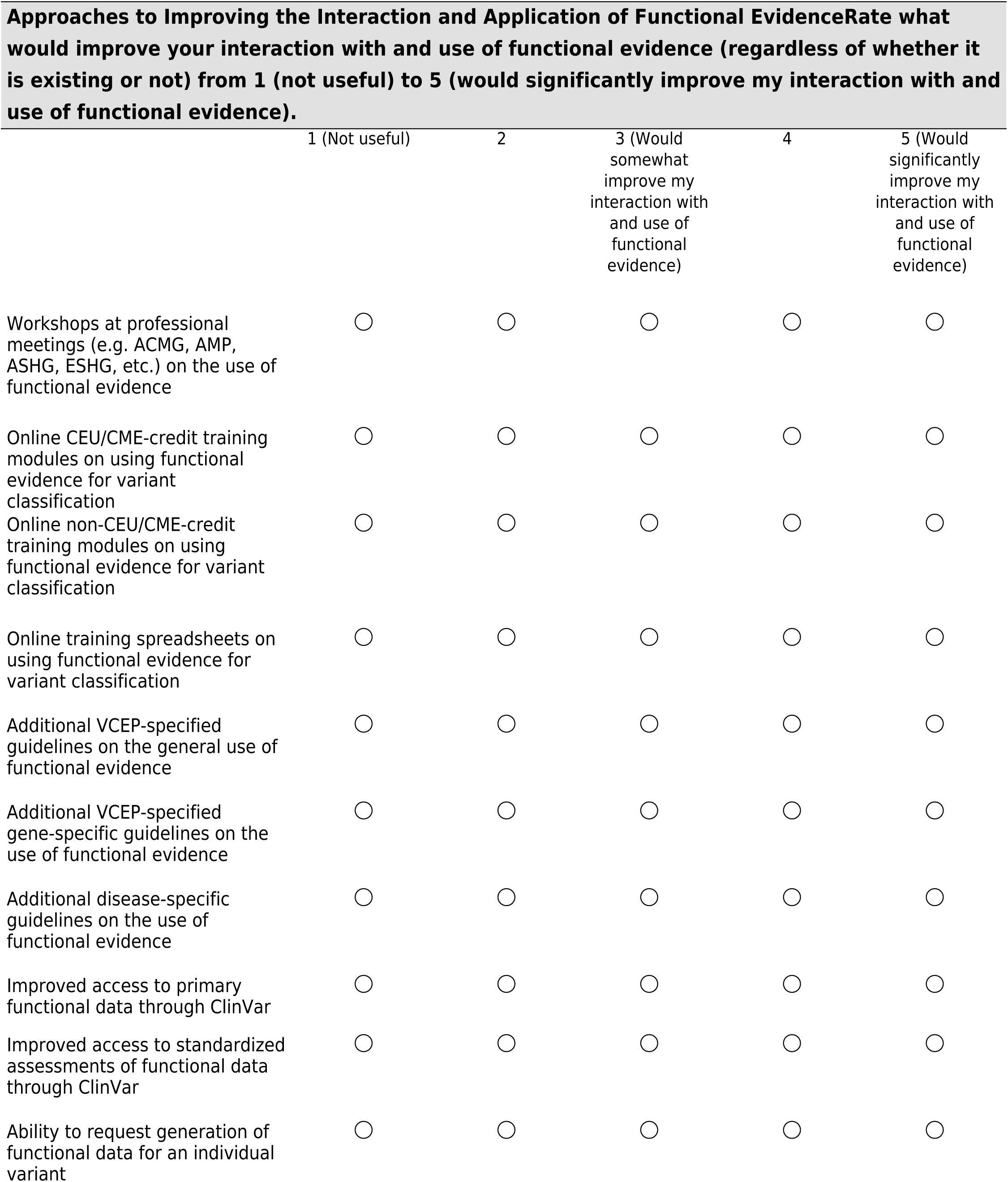
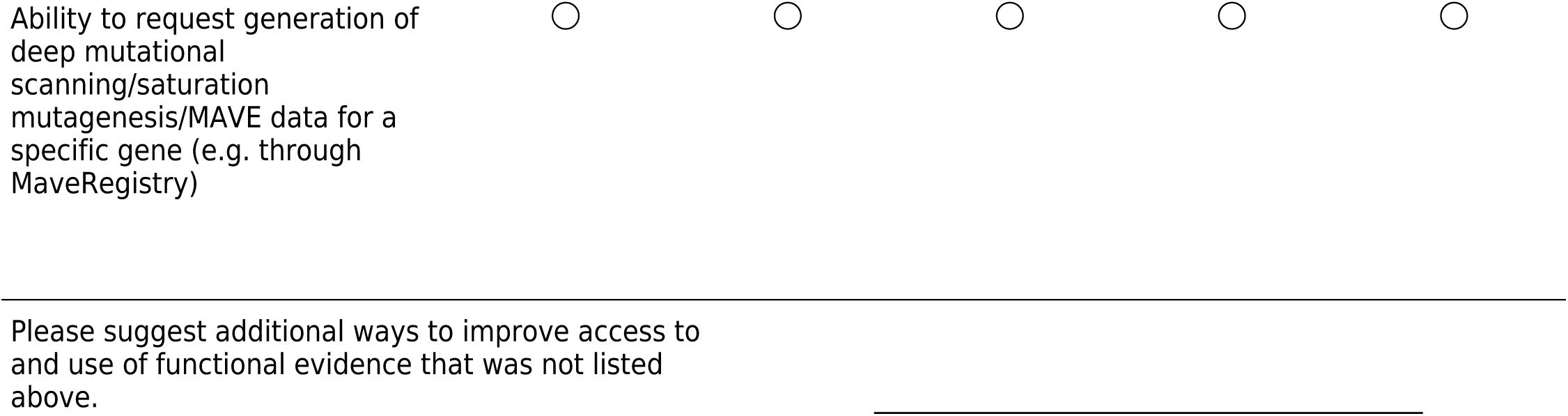

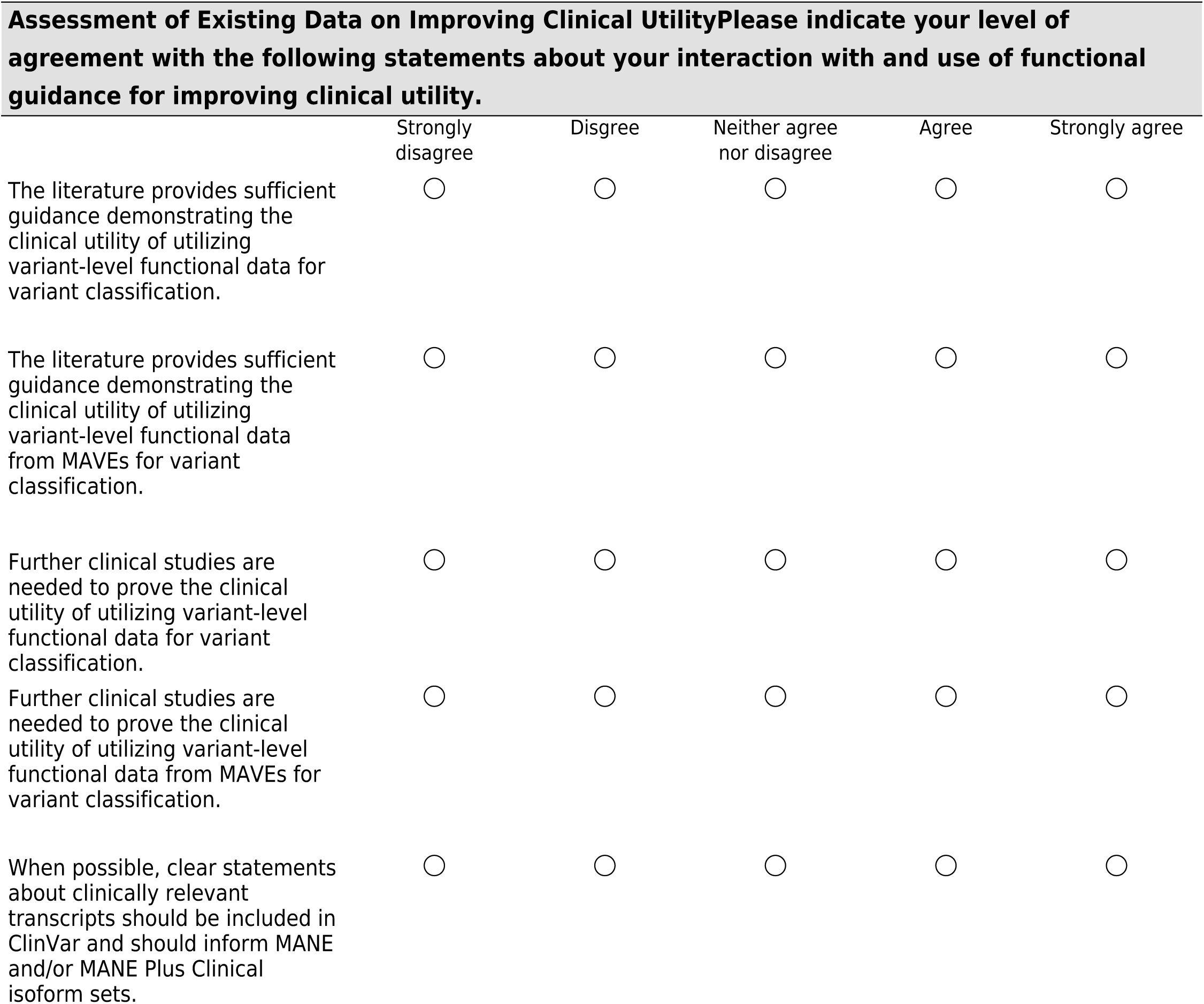

|  | Strongly disagree | Disagree | Neither agree nor disagree | Agree | Strongly agree |
| --- | --- | --- | --- | --- | --- |
| The literature provides sufficient guidance demonstrating the clinical utility of utilizing variant-level functional data for variant classification. | ☐ | ☐ | ☐ | ☐ | ☐ |
| The literature provides sufficient guidance demonstrating the clinical utility of utilizing variant-level functional data from MAVEs for variant classification. | ☐ | ☐ | ☐ | ☐ | ☐ |
| Further clinical studies are needed to prove the clinical utility of utilizing variant-level functional data for variant classification. | ☐ | ☐ | ☐ | ☐ | ☐ |
| Further clinical studies are needed to prove the clinical utility of utilizing variant-level functional data from MAVEs for variant classification. | ☐ | ☐ | ☐ | ☐ | ☐ |
| When possible, clear statements about clinically relevant transcripts should be included in ClinVar and should inform MANE and/or MANE Plus Clinical isoform sets. | ☐ | ☐ | ☐ | ☐ | ☐ |

|  | Strongly disagree | Disagree | Neither agree nor disagree | Agree | Strongly agree |
| --- | --- | --- | --- | --- | --- |
| When classifying variants using gene-level data, I always read the literature that describes this functional data. | ☐ | ☐ | ☐ | ☐ | ☐ |
| Gene-level data is valuable for interpreting pLOF (predicted loss-of-function) variants when variant-specific data is unavailable. | ☐ | ☐ | ☐ | ☐ | ☐ |
| Gene-level experiments provide useful insights for understanding pLOF variants, even when the variants do not exactly match known cases. | ☐ | ☐ | ☐ | ☐ | ☐ |

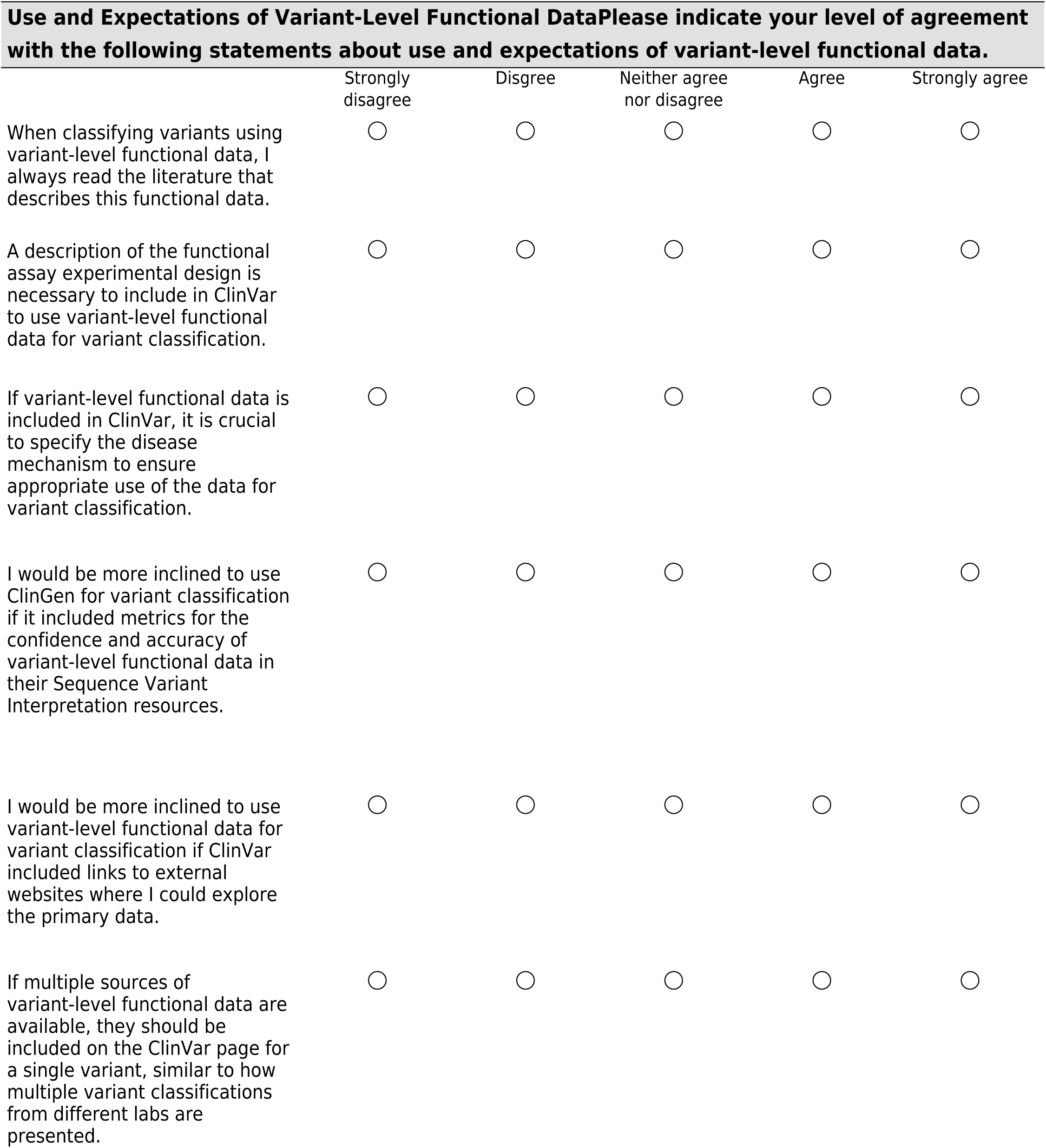
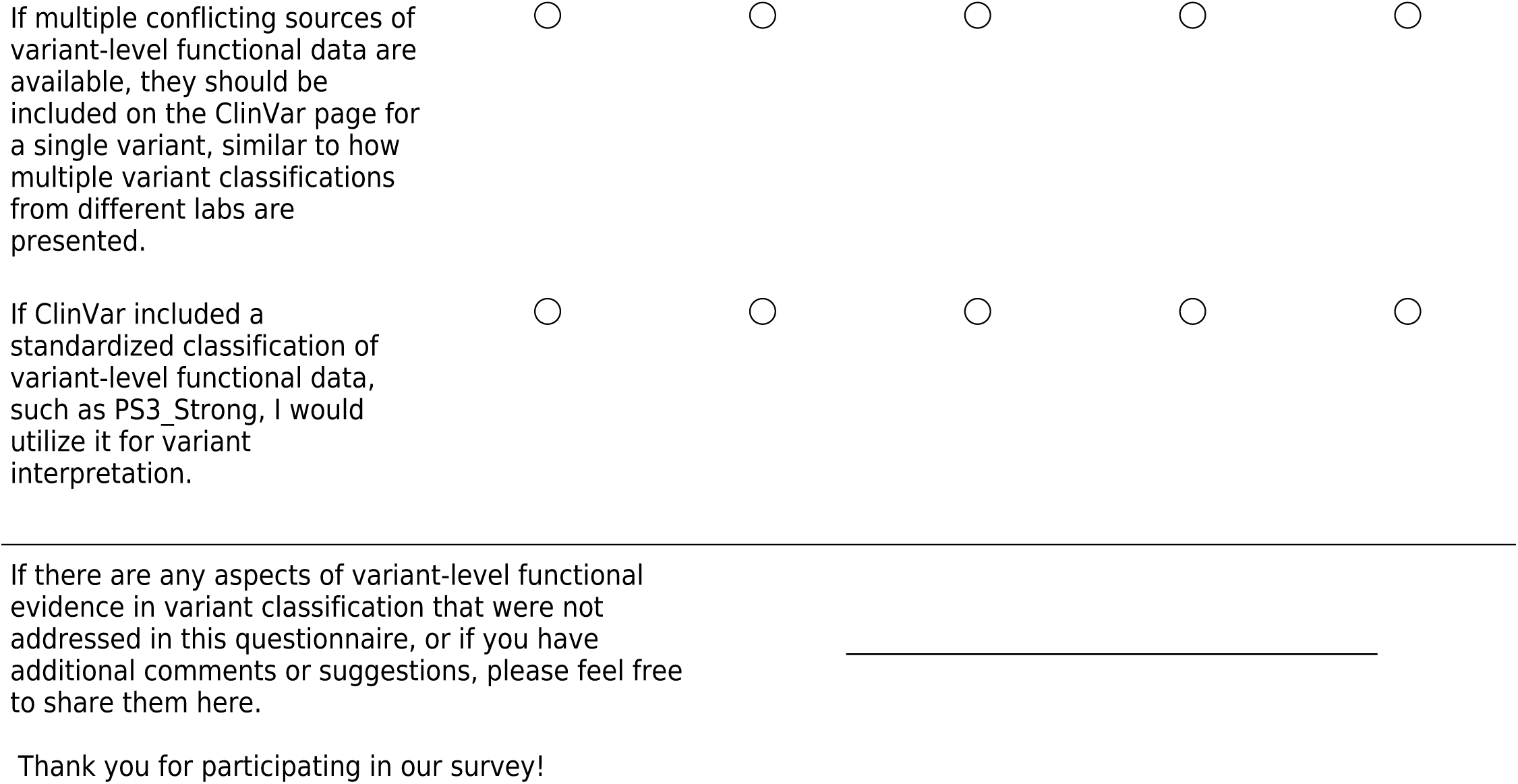

## Additional File 2

**Supplemental Figure 1.**
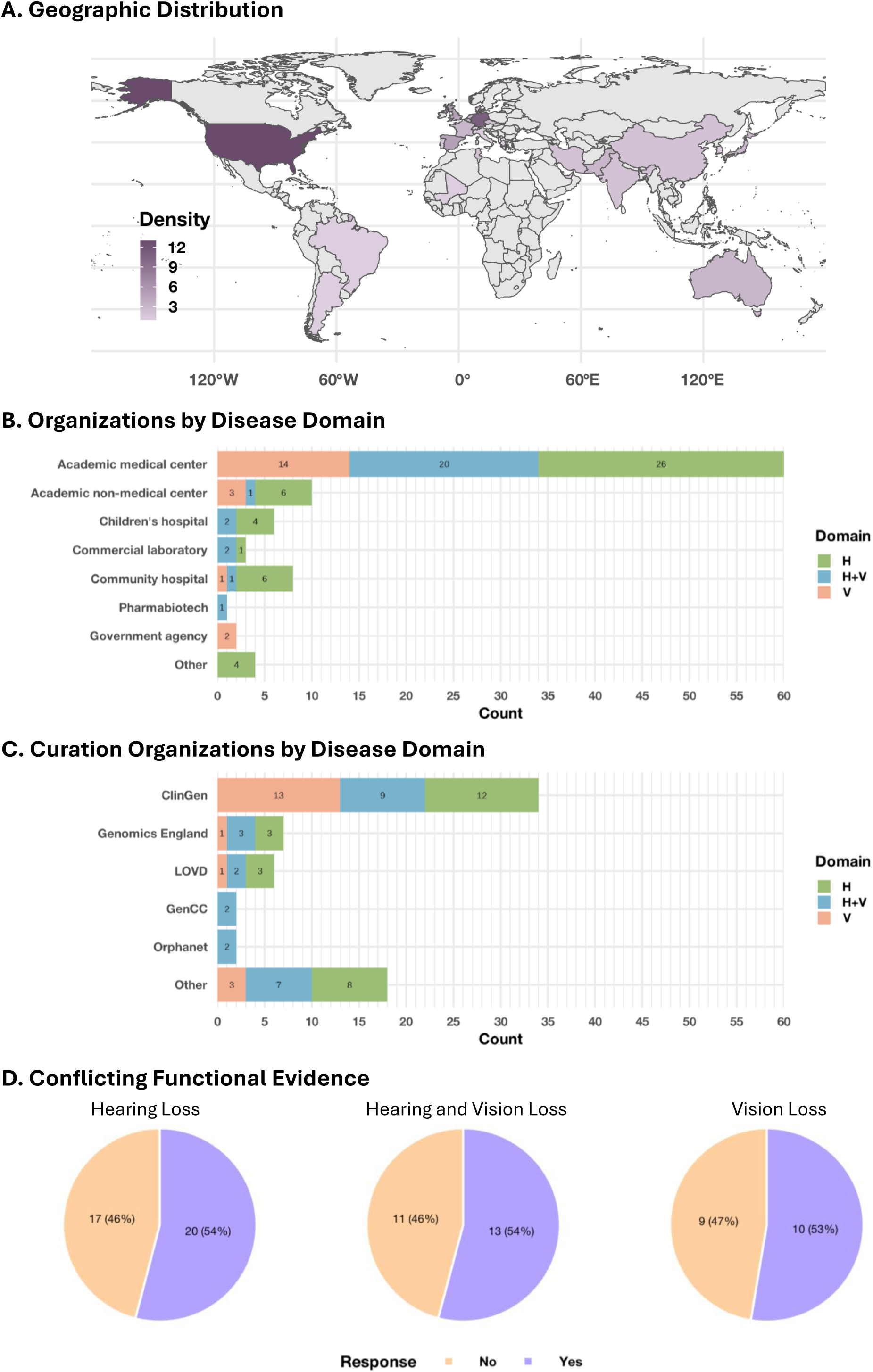
P**a**rticipant **characteristics and experiences with functional evidence.** (**A**) Distribution of respond-ents by organization type. (**B**) Distribution of respondents by curation organization type. (**C**) Geographic distribution of respond-ents. (**D**) Proportion reporting conflicting functional evidence during variant classification.

**Supplemental Figure 2.**
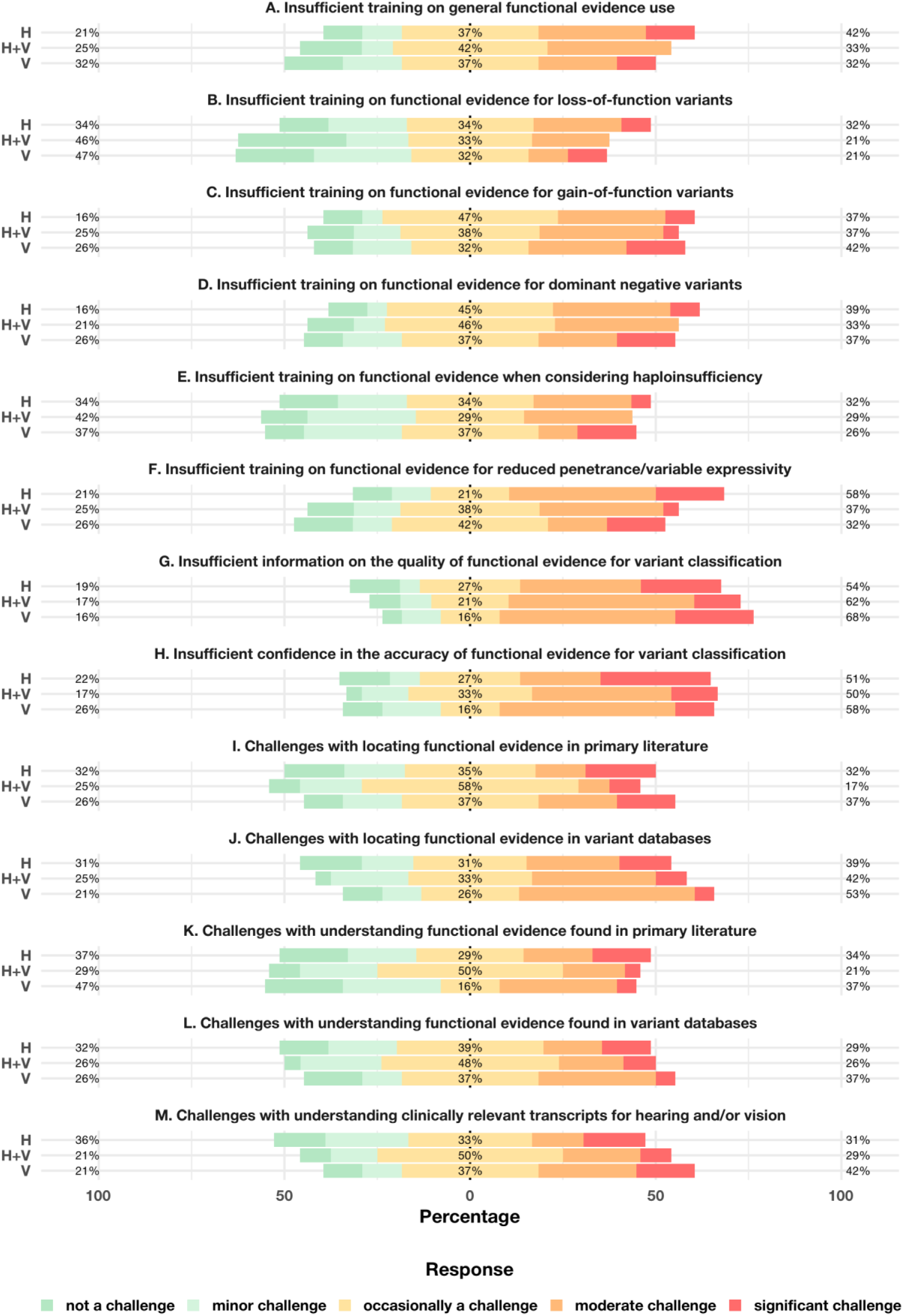
C**h**allenges **in using functional evidence for variant interpretation.** Stacked bar plots display bar-riers to applying functional evidence (**A**-**M**) across disease domains: hearing loss (H), ocular diseases (V), and both domains (H+V). Responses were collected on a five-point Likert scale, ranging from not a challenge (green) to very confident (red), and grouped into three percentage categories: (i) not a challenge/minor challenge, (ii) occasionally a challenge, and (iii) moderate challenge/significant challenge. Missing responses (1-3 per item) were not shown.

**Supplemental Figure 3.**
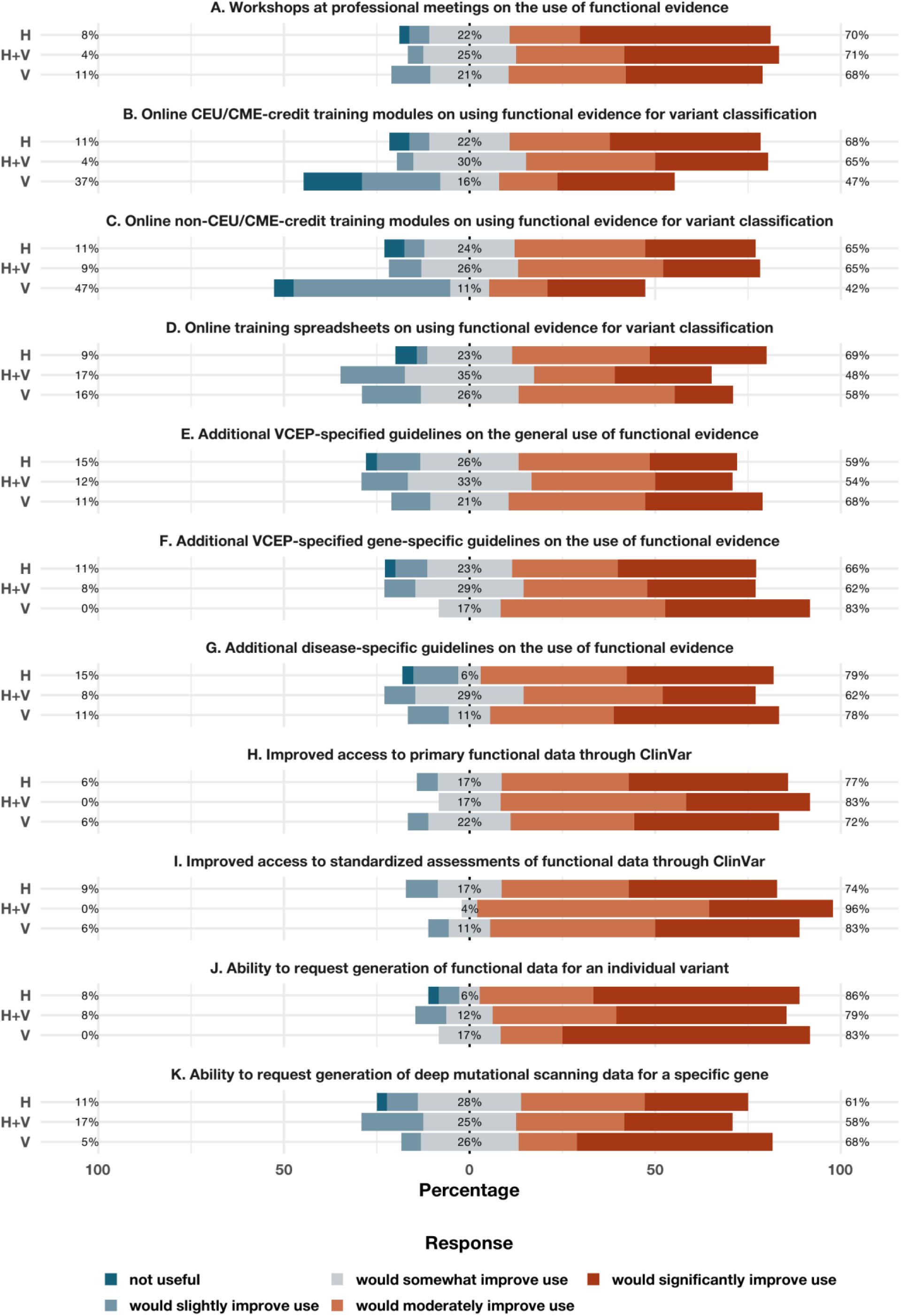
A**p**proaches **to improving the interaction and application of functional evidence.** Stacked bar plots display usefulness of various strategies to enhance the use of functional evidence (**A**-**K**) across disease domains: hearing loss (H), ocular diseases (V), and both domains (H+V). Responses were collected on a five-point Likert scale and grouped into three percentage categories: (i) not useful/would slightly improve use (blue), (ii) would somewhat improve use (grey), and (iii) would moderately improve use/would significantly improve use (red). Missing responses (2-7 per item) were not shown.

**Supplemental Figure 4.**
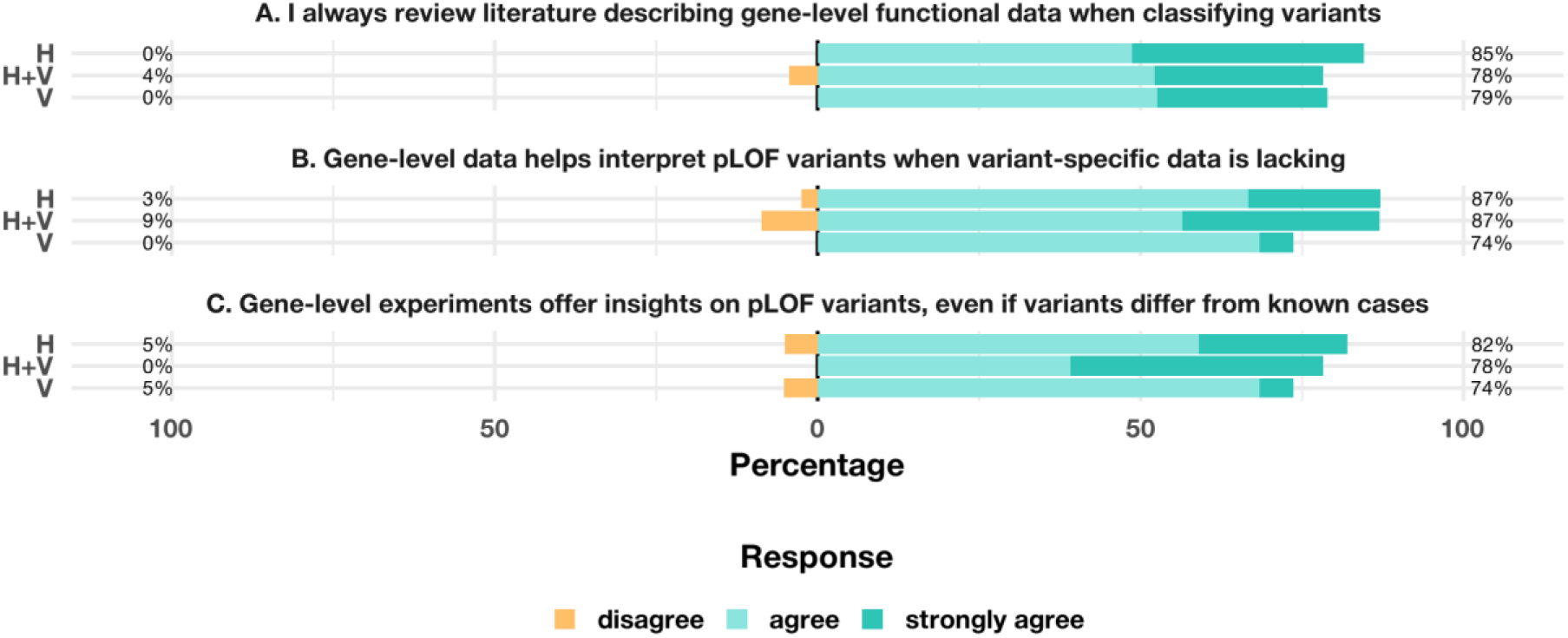
U**s**e **and expectations of gene-level functional data.** Stacked bar plots display agreement with statements regarding interaction with gene-level functional data (**A**-**C**) across disease domains: hearing loss (H), ocular dis-eases (V), and both domains (H+V). Responses were collected on a five-point Likert scale, ranging from strongly disagree (or-ange) to strongly agree (cyan), and grouped into two percentage categories: (i) strongly disagree/disagree, and (ii) agree/strongly agree. Missing responses (1 per item) were not shown. Neutral responses are excluded from display.

**Supplemental Figure 5.**
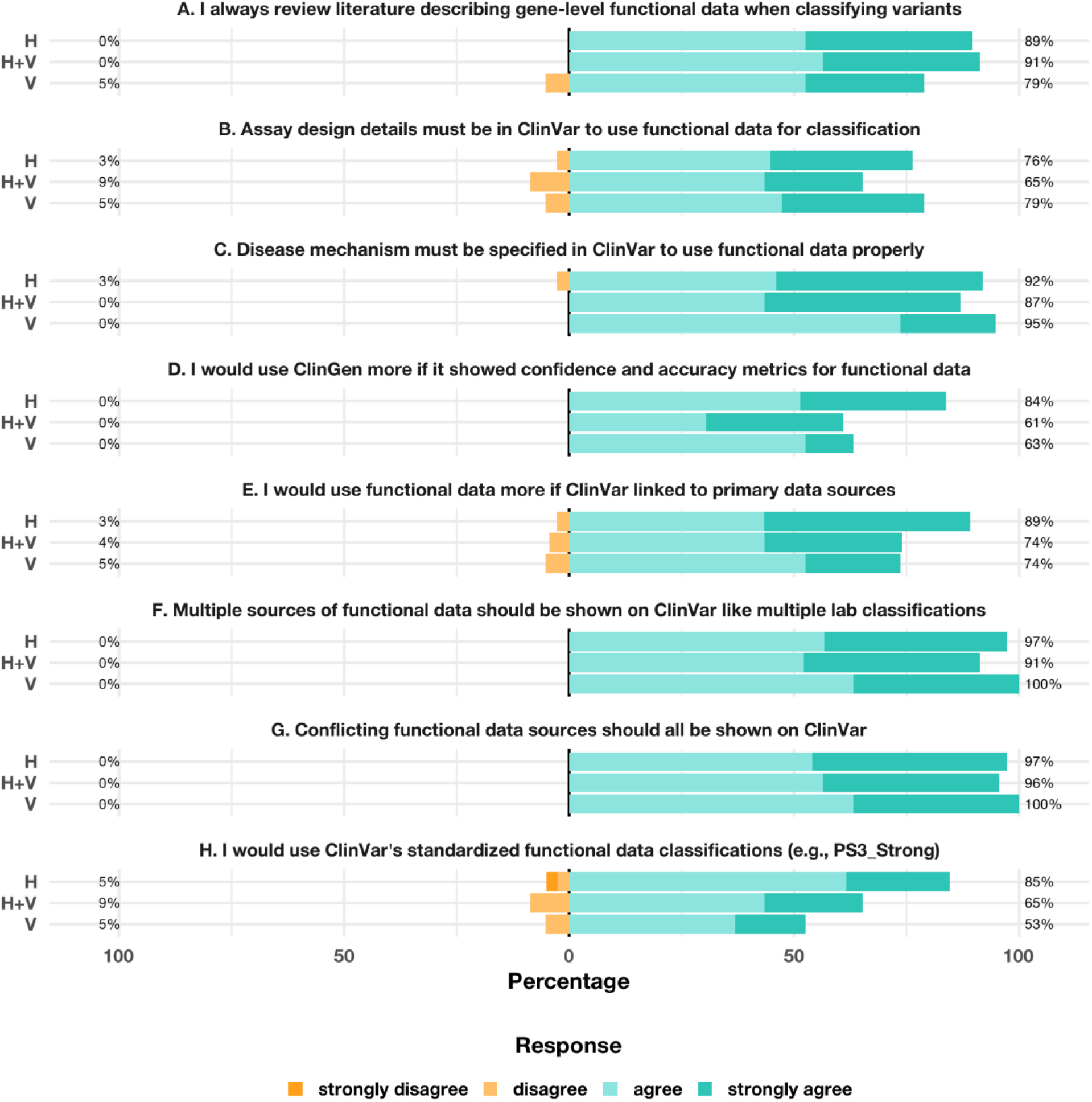
U**s**e **and expectations of variant-level functional data.** Stacked bar plots display agreement with statements regarding interaction with variant-level functional data (**A**-**C**) across disease domains: hearing loss (H), ocular diseases (V), and both domains (H+V). Responses were collected on a five-point Likert scale, ranging from strongly disagree (orange) to strongly agree (cyan), and grouped into two percentage categories: (i) strongly disagree/disagree, and (ii) agree/strongly agree. Missing responses (1-3 per item) were not shown. Neutral responses are excluded from display.

